# Multivariate Modeling of Direct and Proxy GWAS Indicates Substantial Common Variant Heritability of Alzheimer’s Disease

**DOI:** 10.1101/2021.05.06.21256747

**Authors:** Javier de la Fuente, Andrew D. Grotzinger, Riccardo E. Marioni, Michel G. Nivard, Elliot M. Tucker-Drob

## Abstract

Genome-wide association studies (GWAS) of proxy-phenotypes using family history of disease (GWAX) substantially boost power for genetic discovery when combined with direct case-control GWAS, most prominently in the context of Alzheimer’s Disease (AD). However, despite twin study heritability estimates of approximately 60%, recent SNP-based estimates of common variant heritability of AD from meta-analyzed GWAS-GWAX data have been particularly low (2.5%), calling into question the prospects of continued progress in AD genetics. We demonstrate that commonly used approaches for combining GWAX and GWAS data produce dramatic underestimates of heritability, and we introduce a multivariate method for estimating individual SNP effects and recovering unbiased estimates of SNP heritability when combining GWAS and GWAX summary data. We estimate the SNP heritability of Clinical AD diagnoses excluding the *APOE* region at ∼6-10%, with the corresponding estimate for biological AD (based on prevalence rate estimates from recently published molecular imaging data) as high as ∼20%. Common variant risk for AD appears to represent a very strong effect of *APOE* superimposed upon a relatively diffuse polygenic signal that is distributed across the genome.

## Introduction

Genome-wide association studies (GWAS) of proxy-phenotypes using family history of disease (GWAX) can substantially boost power when combined with traditional case-control GWAS across a range of disease traits^1,2^. The benefits of GWAX for enhancing GWAS discovery have been particularly pronounced in the context of late-onset Alzheimer’s Disease (AD)^3–5^. AD is a progressive neurocognitive disorder of aging that is typically marked by a decades-long prodromal period of accumulation of amyloid-predominant neuritic plaques, tau-predominant neurofibrillary tangles, and cortical atrophy, which give rise to a later clinical phase marked by precipitous cognitive decline, impaired activities of daily living, and eventual mortality^6–8^. Recent meta-analyses combining GWAX with direct GWAS have expanded the number of AD-relevant loci far beyond the well-established *APOE* variant, to 75 loci in total^5^. However, despite a twin-based heritability estimates of approximately 60%^9^, these studies indicate very low common variant SNP heritability estimates after excluding the *APOE* region, with the most recently reported estimate of 2.5% on the liability scale coming from the application of LD Score Regression (LDSC) to the largest combined GWAX-GWAS meta-analysis of AD to date^4^. This estimate is noticeably lower than those obtained from the application of both LDSC and raw data-based methods in earlier studies that have only included direct case-control designs, albeit in smaller samples^10–12^. If valid, this low SNP heritability estimate may call into question the prospects of continued progress in genetic discovery for AD within the spectrum of common variants.

Here, we identify several sources of downward bias in commonly used approaches for the estimation of individual SNP effects and SNP heritability from GWAX summary data. We show that, under scenarios in which the assumptions of the standard GWAX model are met, commonly used approaches for combining GWAX and GWAS data produce dramatic underestimates of SNP heritability, and we provide simple corrections to remove such biases. We go on to consider a range of common violations of the standard GWAX model that may produce further downward bias, and we develop a more flexible data-driven multivariate framework for the accurate estimation of SNP heritability from GWAS and GWAX summary data in the presence of such violations. Our approach produces consistent estimates of SNP heritability both in best-case scenarios in which standard approaches fail, and in more complex scenarios in which assumptions of the standard GWAX model are violated. Our model relaxes the assumption that the phenotype has been measured with equal fidelity in cases and proxy cases, and that the genetic relationship between genotyped individuals and their affected and unaffected family members is known with certainty.

Using recently released summary GWAS data from the International Genomics of Alzheimer’s Project (IGAP)^10^ and GWAX data from UK Biobank^3^ our multivariate method yields substantially increased heritability estimates for clinical AD relative to the recently reported estimates from naïve meta-analysis of GWAS and GWAX data. Because the steep age-graded nature of AD risk and the insidious “silent period” of AD progression renders identification of a single AD prevalence rate difficult, we examine how estimates of AD heritability are affected by various assumptions about prevalence rate that are informed by epidemiological and molecular imaging data. We find that, in contrast to the most recently reported SNP heritability estimate of 2.5% for AD when excluding the *APOE* region, the value for clinical AD is likely to between approximately 6% and 10%, and the value for biological AD (as defined by pathological levels of amyloid and tau in the brain) may be as much as 20%. Cumulative analysis of local SNP heritability indicates that while the *APOE* locus accounts for approximately one quarter of common variant SNP heritability, the remaining common variant SNP heritability appears to represent polygenic signal that is relatively diffusely distributed across the genome.

## Results

### Overview of Multivariate Method

Recent large scale meta-analyses combining direct-GWAS and GWAX data have used either inverse variance weighted meta-analysis of regression coefficients^3,13^ or sample size weighted meta-analysis of Z statistics^4,14^. For the inverse variance approach, the standard correction factor for GWAX of the phenotype of a single first degree relative using the offspring genotype is to multiply regression coefficients and their SEs by 2.0 to correct for 50% attenuation due to 50% genetically relatedness. A similar correction to the Z statistics approach is possible (section S3 of the Supplementary Note) but not typically made. However, even with such correction, estimation of SNP heritability from the summary statistics produced by either method produces estimates that are severely biased, unless the sample size input is further corrected (section S4 of the Supplementary Note). Moreover, GWAX estimates will be attenuated by more than the 50% assumed by the standard correction under a wide range of circumstances such as those in which: a proportion of genotyped individuals report on the phenotypes of their step or adoptive parents (such that average genetic relatedness of phenotyped and genotyped individuals falls below 50%), when individuals are not well-informed about, misremember or confuse their parents’ phenotype or disease status (such that heritability of the GWAX phenotype is attenuated, or contaminated by other heritable phenotypes), or when the average diagnostic quality or criteria differ for proxy reports of historical disease status and direct GWAS of carefully screened case-control sample (such that heritability of the GWAX phenotype is attenuated). We develop a multivariate model that directly estimates the appropriate correction empirically from the GWAX and direct GWAS summary data in order to produce unbiased estimates of SNP heritability and SNP effects without manual correction of effect size estimates, standard errors, or sample sizes.

Our multivariate model (Supplementary Note Section S5 and Supplementary Figure S1) integrates summary data from three sources: direct GWAS, maternal GWAX, and paternal GWAX. In the model, the total genetic propensity toward AD risk is represented as a latent factor, *F*, that is specified to affect the direct GWAS phenotype and two GWAX phenotypes according to the following system of regression equations

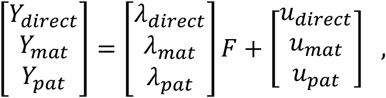

where the *λ* coefficients relate *F* to measured phenotypes *y*, and the *u* terms are residual genetic propensities toward each of the measured phenotypes that are independent of *F*, and uncorrelated with one another and with *F*. We specify the model with the minimal identification constraint that *λ*_*direct*_ = 1 such that *F* takes on the scale of the direct GWAS phenotype, and 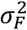 can be interpreted as an unbiased estimate of the SNP heritability of the meta-analyzed phenotype in the direct GWAS metric. Note that the standard GWAX approach^1^ implicitly treats *λ*_*Mat*_ = *λ*_*Pat*_ = 5 and 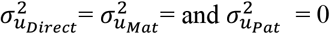. Thus, under our more flexible multivariate parameterization in which these terms are freely estimated, the departure of *λ*_*Mat*_ and *λ*_*Pat*_ from.5 and departure of the 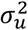 terms from 0 indicate departure of the empirical data from the standard GWAX assumptions.

To estimate meta-analytic summary statistics using this multivariate model (section S6 of the Supplementary Note), we expand it to include the effect of an individual genetic variant, *x*, on the *F* as follows

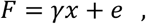

where ϒ is an unstandardized regression coefficient and *e* is a residual. Models are estimated in Genomic SEM^15^ using a two-stage approach. In the first stage, the empirical liability-scale genetic covariance matrix and its sampling covariance matrix are estimated. In the second stage, the model is fit to the matrices using the diagonally weighted least squares (WLS) fit function with sandwich correction, as described in Grotzinger et al.^15^

### Simulation Results

We simulated genome-wide summary statistics for direct GWAS and maternal and paternal GWAX, independently manipulating 1) the sample size for direct GWAS and maternal and paternal GWAX summary statistics, and 2) the relative effect sizes for maternal and paternal GWAX compared to the direct GWAS (i.e. the attenuation coefficients, *λ*). We compared the Genomic SEM approach to the conventional uncorrected approach (directly reflective of those taken by Jansen et al.^14^ and Wightman et al.^4^) and the conventional corrected approach (directly reflective of those taken by Marioni et al.^3^, and Bellenguez et al.^5^)

The top row of Figure 1 presents simulation results with respect to SNP heritability estimates from conditions in which sample size is held constant and the *λ* coefficients vary. Complete results for recovery of SNP heritability across all conditions are reported in Supplementary Table S2 and displayed in Supplementary Figure S2. The multivariate Genomic SEM approach provided essentially unbiased heritability estimates 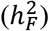 across all conditions, outperforming the conventional (uncorrected) approach in all conditions, and outperforming the conventional (uncorrected) and conventional (standard correction) approaches in conditions in which the assumptions of the simple GWAX model did not hold (Percent Bias Error, %BE, range for conventional [uncorrected] approach: 59.7%-89.0%; %BE range for conventional [standard correction]: 6.6%-55.9%. As expected, the conventional approach with the standard correction performed similarly to the Genomic SEM approach when the standard assumption that *λ*_*Mat*_ = *λ*_*Pat*_ =.5 held. Bias in the conventional approaches was related to the extent to which the population values of *λ*_*Mat*_ and *λ*_*Pat*_ deviated from.5 and the sample size of the relative contribution of the GWAXs to the total sample size.

**Figure 1.**
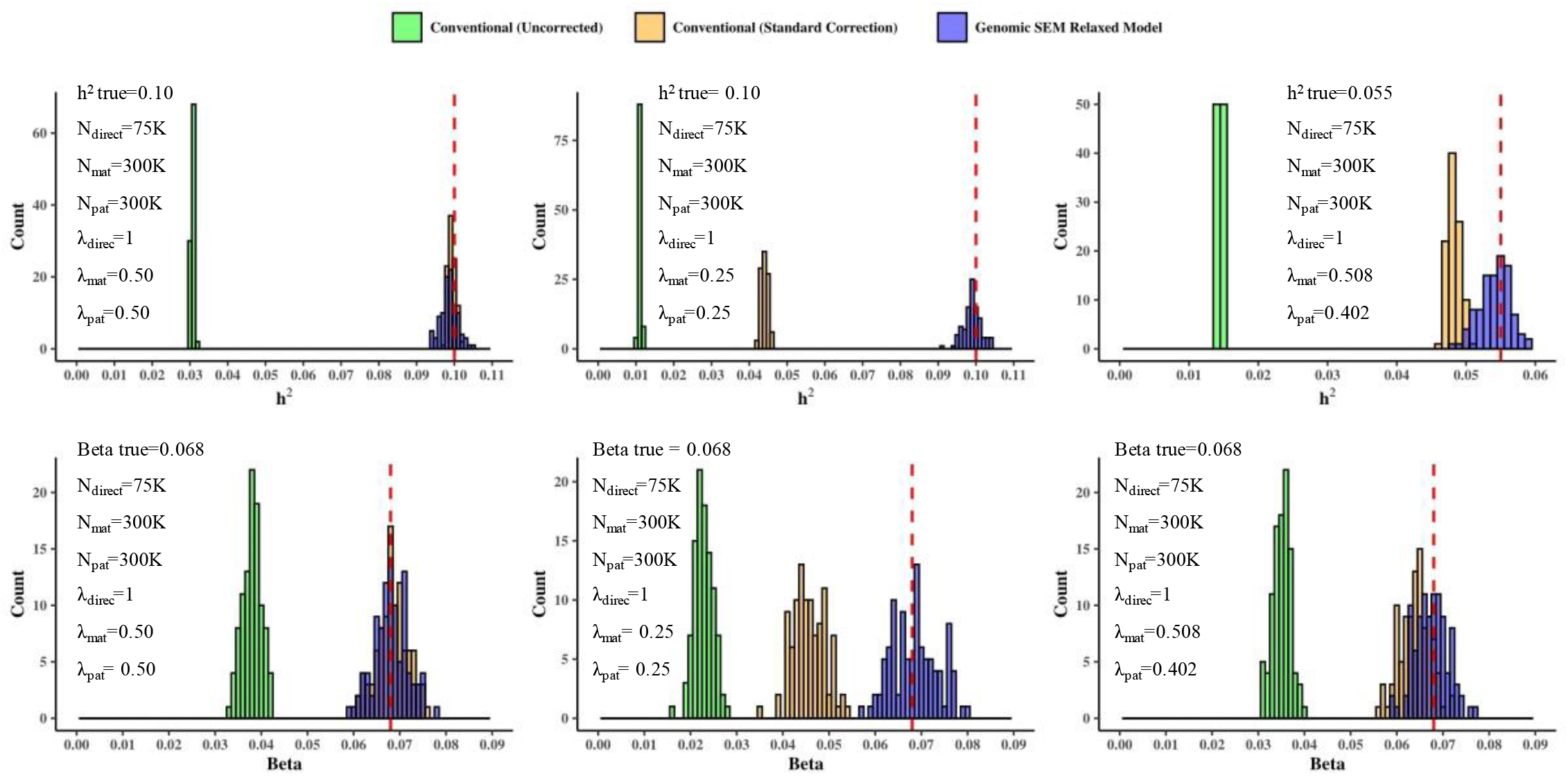
Simulation Results. Distribution of SNP heritability estimates (top row) and individual SNP effects (bottom row) for Conventional (Uncorrected), Conventional (Standard Correction), and Multivariate model for combining GWAS and GWAX estimates across a range of attenuation coefficients for the GWAX in the population. The vertical dashed red lines indicate the true parameter value in the population. Complete simulation results for all conditions are reported in Supplementary Tables S2 and S3 and displayed in Supplementary Figures S2 and S3.

The bottom row of Figure 1 presents simulation results with respect to individual SNP effects for conditions in which sample size is held constant and the *λ* coefficients vary. Complete results for recovery of individual SNP effects across conditions are provided in Supplementary Table S3 and Supplementary Figures S3. Results for the recovery of individual SNP effects were consistent with those for the recovery of heritability. Our multivariate Genomic SEM approach exhibited consistently unbiased performance across all conditions (all %BE <1%), outperforming the uncorrected approach in all conditions and outperforming the corrected approach in those conditions for which the simple GWAX model did not hold (%BE range: 36.59%-66.48% uncorrected; %BE range: 3.46%-32.84% corrected). When the simple GWAX model assumptions held, the Genomic SEM approach and the conventional approach with the standard correction were both unbiased, but the conventional approach without correction remained biased. Bias in the conventional approaches was related to the extent to which the population values of *λ*_*Mat*_ and *λ*_*Pat*_ deviated from.5 and the relative contribution of the GWAXs to the total sample size.

### Multivariate Model of Direct GWAS and GWAX of Alzheimer’s Disease

We applied our multivariate model to empirical summary data from the direct case-control GWAS of AD in IGAP^10^ and GWAX of maternal and paternal AD in UK Biobank^3^. Key descriptive statistics for these three contributing datasets are reported in the top portion of Table 1, and multi-trait LDSC results (cross trait intercepts and genetic correlations) are provided in Supplemental Figure S4. Note that all LDSC estimates reported in Table 1 and Supplementary Figure S4 are based on common variants (MAF ≥.01) outside of the MHC and *APOE* regions, using the AD population prevalence rate of 4.3% assumed by Jansen et al.^14^. Genetic correlations were close to 1.0 between IGAP and maternal AD (rG = 0.915, SE = 0.153, *p* < 0.001) and paternal AD (rG = 0.842, SE = 0.206, *p* < 0.001) in UKB. The genetic correlation between maternal and paternal AD was also very high (rG = 0.815, SE = 0.253, *p* = 0.001). In the bottom portion of Table 1 we provide key descriptive of summary statistics from our multivariate meta-analysis and from two recent meta-analyses that have combined GWAS and GWAX AD data^3,14^. A complete description of findings of our multivariate meta-analysis with respect to individual SNP effects can be found in in section SR1 of the Supplementary Results, and Supplementary Figures S5 and S6.

**Table 1.**
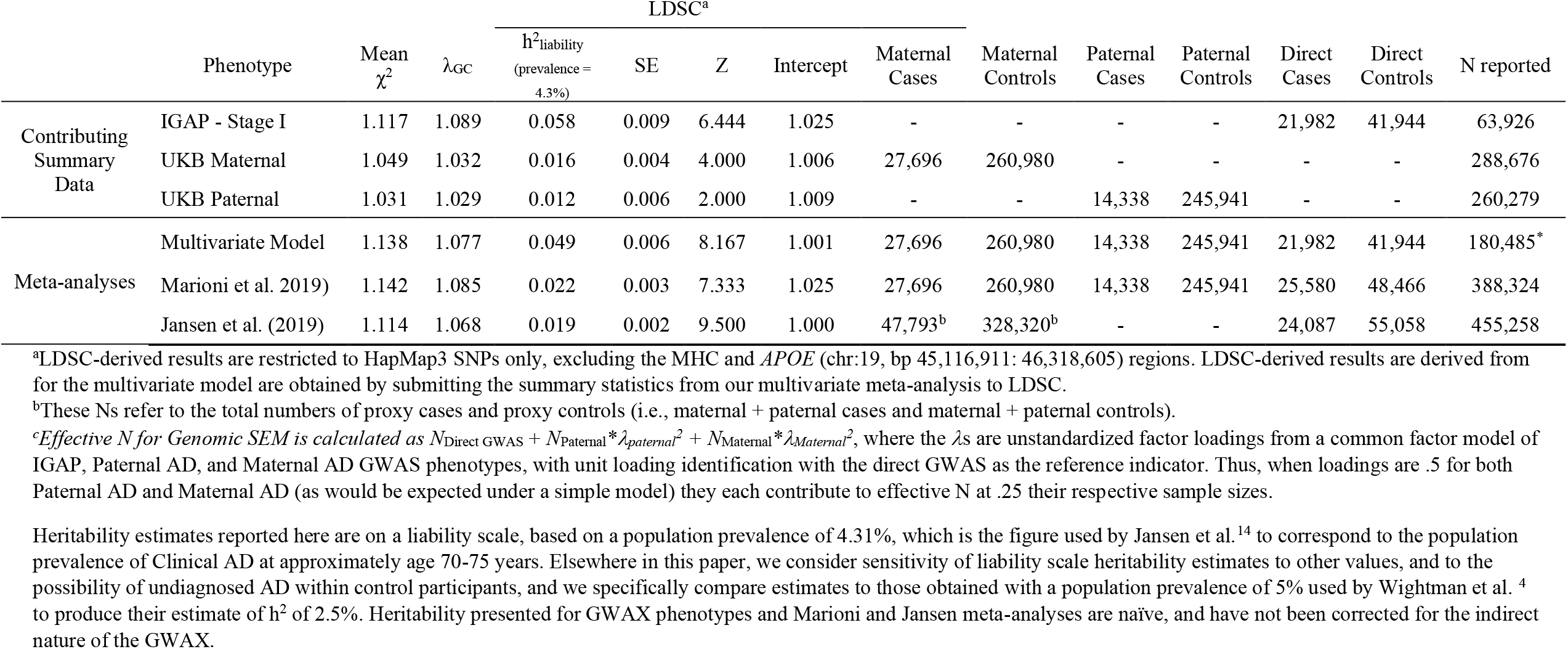
Descriptive statistics for contributing cohorts and previous meta-analyses of case-control and proxy-phenotypes of Alzheimer’s disease and LDSC output.

Estimates from the full multivariate model of AD are displayed in Figure 2. In the model itself, the liability-scale SNP heritability estimate was 5.5% (SE=2.4%), over double that estimated by application of LDSC to the summary statistics from the other two recent AD meta-analyses, using their reported sample sizes. A version of the model that constrained residual variance of the direct GWAS to 0 (reflecting the assumption that the direct GWAS does not contain ancillary genetic signal unrelated to AD) did not produce a significant decrement in model fit (χ^2^(1) =.021, p =.89) and produced a very similar SNP heritability estimate (5. 8%) with a substantially smaller standard error (SE = 1%). Applying LDSC to GWAS summary statistics produced by our multivariate method using the effective sample size derived from the attenuation estimates from the model also produced a similar SNP heritability estimate (4.9%, SE=.6%; Table 1). In the full model, the unstandardized loading for maternal AD (*λ*_*Mat*_ = .508)was nearly identical to the expectation under the standard GWAX model (*λ*_*Mat*_ =) Conversely, the unstandardized loading for paternal AD (*λ*_*Pat*_= 402) reflected approximately 20% attenuation of regression effects (i.e.[.5-.402]/.5) and approximately 35% attenuation of R^2^ and h^2^ estimates ([.5^2^-.402^2^]/[.5^2^]) relative to the expectation under the standard model (*λ*_*Pat*_= 5). Allowing the *λ*_*Mat*_ and *λ*_*Pat*_ parameters to be freely estimated, can avoid the potential for bias stemming from violations of the standard assumption, in this case particularly for paternal effects, when estimating genome wide meta-analytic summary statistics. We note that the heritability estimates reported in this section are based on an assumed AD population prevalence rate of 4.3%, as per the LDSC analyses by Jansen et al.^14^. Below, we consider sensitivity of the heritability estimate to different assumptions regarding the sample and population prevalence rates of AD.

**Figure 2.**
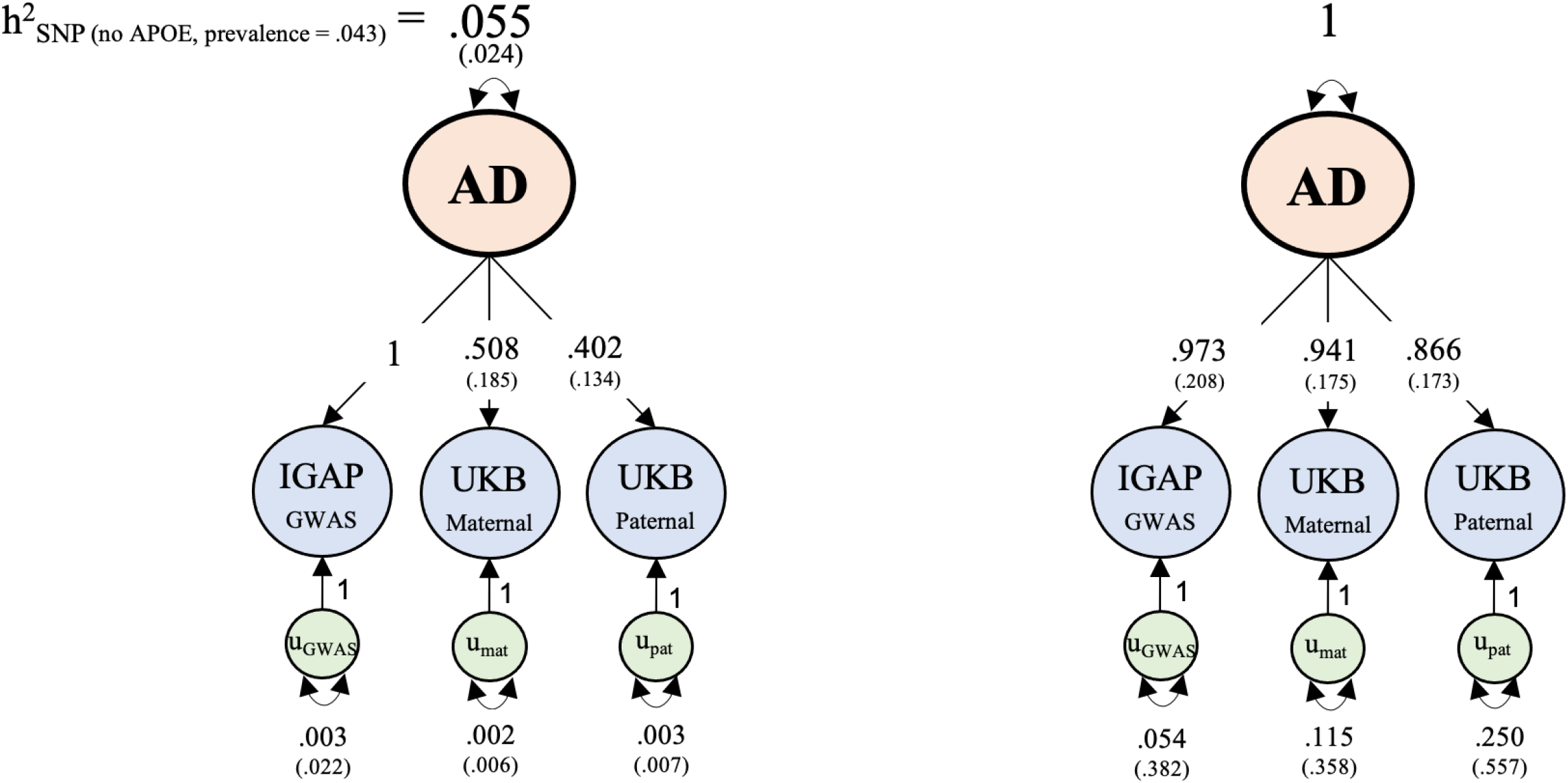
Unstandardized (left) and standardized (right) empirical results from multivariate genetic analysis of Alzheimer’s disease. The liability-scale SNP heritability estimate of 5.5% is on the scale of the direct GWAS, and is for common variants (MAF ≥.01) outside of the MHC and *APOE* regions, using the AD population prevalence of 4.3% assumed by Jansen et al.^14^

### Liability Scale Heritability across a Range of Assumptions

Heritability estimates for case-control traits, such as AD, are based on a liability threshold model, which assumes a continuously distributed liability toward the binary phenotype in the population. Estimates of liability-scale SNP heritability are sensitive to assumptions about the lifetime prevalence of the disorder in the population and the extent to which unaffected individuals have been successfully screened for the disorder (see Method). When population prevalence rates are high or when control participants are unscreened, differences in allele frequencies between cases and controls represent less extreme comparisons along regions of the liability distribution. In such circumstances, the inferred liability scale heritability is higher than would be inferred from the same case-control difference in allele frequencies for a disorder with a lower population prevalence or when control participants have been carefully screened. This issue is particularly germane to the study of AD, a disorder whose: (i) clinical prevalence rate increases from less than 1% in middle adulthood to approximately 30% by old age^16^, (ii) that is known to go undetected at high rates for decades prior to diagnosis due to ancillary factors (e.g. Educational attainment) unrelated to biological severity^17^, and (iii) whose pathophysiological basis may be more than twice as prevalent as its clinical diagnosis at any given age^18^.

To gauge the effects of assumptions regarding population prevalence on our SNP heritability estimate for AD, we varied the assumed population prevalence rate. We provide rough age equivalents of these prevalence rates based on published epidemiological data for Clinical AD^16^, assuming that control participants were appropriately screened. We refer to this estimate as the estimate of heritability of Clinical AD, in that this estimate does not account for undetected biological AD that may be undetected in control participants. Estimates from the full multivariate model are displayed in the left panel of Figure 3 (Estimates from the application of LDSC to the summary statistics from our multivariate meta-analysis were very similar; Figure S7). Heritability estimates increase as the population prevalence rate increases, reaching approximately h^2^ = 10% at prevalence = 30%. We note that Wightman et al.^4^, estimate a heritability of 2.5% assuming a population prevalence rate of 5%. At this same prevalence rate, the estimate from our full multivariate model is over double (liability h^2^ = 5.8%) that of Wightman et al. This is likely a reflection of the fact that Wightman et al. do not appear to make any correction to their LDSC estimate to account for any attenuation in the GWAX estimates due to their indirect nature.

**Figure 3.**
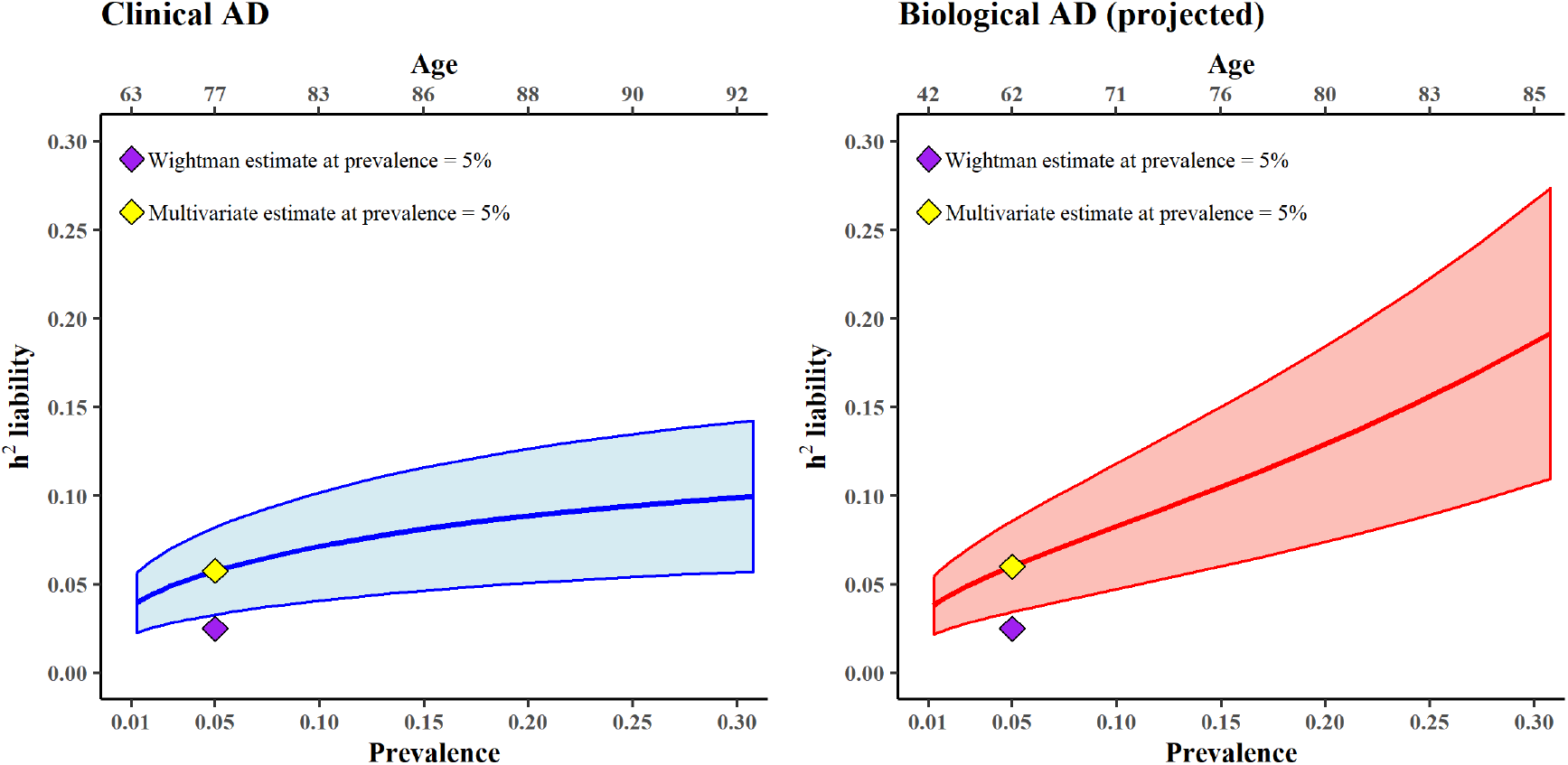
Estimated common variant SNP heritability of AD (outside of the MHC and *APOE* regions) according to different assumptions regarding the population prevalence of AD and biological AD. We provide rough approximations of the age equivalences of each prevalence rate on the top x axis. The purple diamond represents the estimate of 2.5% by Wightman et al.^4^, which was based on an assumed population prevalence rate of 5%. The yellow diamond represents the estimate from the multivariate model introduced here, using the same assumed population prevalence rate of 5% (Clinical AD h^2^ = 0.058; Biological AD h^2^ = 0.06). The shading area around the line reflects +/-1 SE of the h^2^ estimate. The steeper shift in the SNP heritability of AD for biological AD compared to clinical AD as a function of population prevalence stems from the correction for undetected biological AD within control participants who primarily only been screened for clinical AD. Thus, as the assumed prevalence rate of biological AD increases, the extent of case contamination in control participants increases, and the correction for undetected AD in control participants produces more dramatic increases in the projected heritability.

We also estimated SNP heritability (for common variants outside of the MHC and *APOE* regions) across a range of different assumed prevalence rates of biological AD, employing a correction for case contamination (see Method) in control participants (who were primarily screened for Clinical AD but not AD pathology) and provide rough age equivalents of these prevalence rates for biological AD (Alpha+ and Tau+) from recently published positron emission tomography data^18^. We refer to this estimate as the projected estimate of biological AD heritability. Results are displayed in the right panel of Figure 3. Heritability estimates increase steeply as the prevalence rate increases, reaching approximately h^2^ = 20% at a population prevalence of 30%. The steeper shift in the SNP heritability of AD for biological AD compared to clinical AD stems from our correction for undetected biological AD within control participants who have primarily only been screened for clinical AD. As the assumed prevalence rate of biological AD increases, the extent of case contamination in control participants increases, and the correction for undetected AD in control participants produces more dramatic increases in the projected heritability. The validity of this inference relies on the assumptions that individuals with clinical AD diagnoses are representative of the larger set of individuals with biological AD, and do not constitute a subgroup of those with more severe biological AD. This assumption is supported by a large body of work indicating that ancillary factors, such as educational attainment, are associated with clinical AD rates among individuals with equivalent levels of brain pathology^19^.

### Inferring Common Variant Polygenicity from Local SNP Heritability

Our results indicated relatively high common variant SNP heritability of AD outside of the *APOE* locus. To investigate whether this SNP heritability was attributable to a small number of genomic regions, or distributed more evenly across the genome, we used Heritability Estimation from Summary Statistics (HESS)^20^ to estimate local common variant SNP heritability in 1703 approximately independent blocks across the genome using the summary statistics from our multivariate meta-analysis. Following Sinnott-Armstrong et al.^21^, we plot the proportion cumulative heritability of AD liability across the genome in Figure 4. Because HESS allows for negative estimates of local SNP heritability, estimation error is not expected to produce increases in cumulative heritability; only true polygenic signal will produce such increases. It can be seen that heritability accumulates relatively continuously across the entirety of the genome, with a pronounced discontinuity at the AD locus on chromosome 19. This locus accounts for 27.59% of total common variant SNP heritability of AD, indicating that nearly three quarters of common variant SNP heritability is attributable to genetic signal independent of *APOE*. Local SNP heritability in the other loci that were genome-wide significant in the multivariate GWAS was strongly correlated with their GWAS effect sizes (Figure S8) and accounted for an additional 9.60% of the total common variant SNP heritability, leaving 62.82% of the total common variant SNP heritability unexplained by genome-wide significant loci. The observations that heritability accumulates relatively continuously outside of the *APOE* locus and that substantial proportion of SNP heritability remains outside of genome-wide significant loci suggests that genetic risk for AD may be affected by core pathways superimposed on a more diffuse polygenic background. Using the conservative estimate of population prevalence of 4.31%, assumed by Jansen et al.(3), the total heritability of AD on the liability scale as estimated with HESS was 12.36% (8.94% excluding the *APOE* locus). Similar patterns were observed when HESS analyses were limited to GWAS data from IGAP only (Supplemental Figure S9).

**Figure 4.**
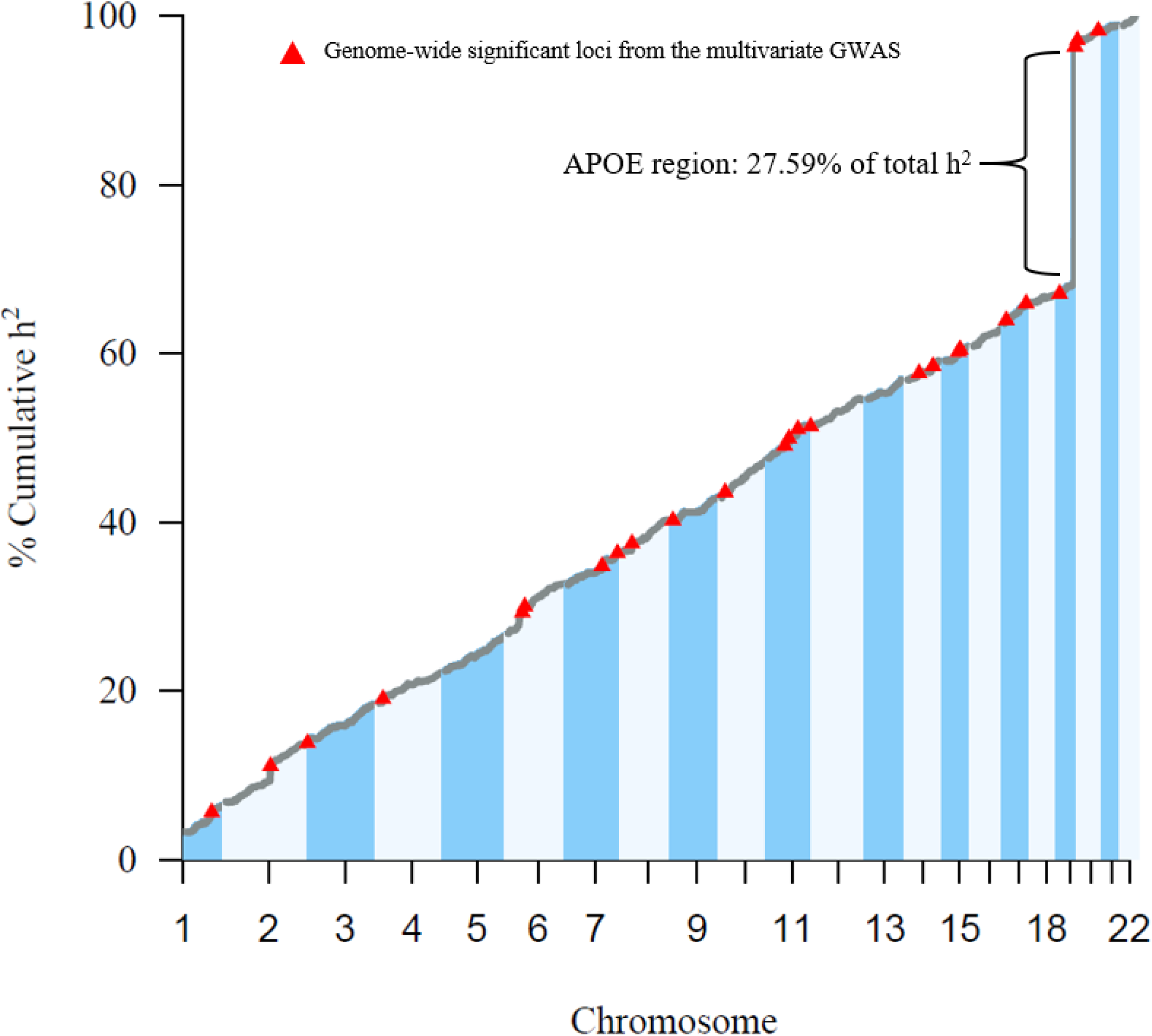
Proportion of cumulative heritability across the genome, as estimated with HESS^20^. Red triangles represent genome-wide significant loci from the multivariate GWAS of AD.

## Discussion

We have introduced and validated a novel multivariate method for the joint analysis of direct GWAS and proxy GWAX summary data. Our method recovers unbiased estimates of common variant SNP heritability and of individual SNP effects under a variety of circumstances in which standard approaches, including those that correct for the indirect nature of the GWAX, dramatically underestimate these quantities. Using recently released GWAS and GWAX summary data for AD, our method estimates the common variant SNP heritability of AD excluding the *APOE* region to be more than double the recent estimate of 2.5% by Wightman et al.^4^, when using same assumptions regarding population prevalence. This estimate rises higher when considering prevalence rates of Clinical AD and Biological AD indicated by epidemiological data.

Analysis of local SNP heritability indicates that over 70% of common variant SNP heritability of AD exists outside of the *APOE* region. We find that this remaining heritability accumulates relatively continuously across the remainder of the autosome, indicating that a portion of genetic risk for AD is characterized by a relatively diffuse polygenic architecture. This pattern resembles that recently observed for three molecular traits whose biological pathways are well-understood^21^. These authors inferred that core gene sets representing proximal biological mechanisms play a sizable role in each trait, but that “most of the SNP-based heritability is driven by a massively polygenic background.” Our results suggest that the same may pertain to the genetic architecture of AD.

Background polygenic architecture may reflect biological mechanisms underlying the pathophysiology of AD or ancillary factors that affect the likelihood of diagnosis of Clinical AD without contributing to disease onset or progression. The strongest candidates for such ancillary factors may be educational attainment and premorbid cognitive functioning, both of which longitudinal research indicates are not related to the onset or rate of cognitive decline but are related to the likelihood that declines will be detected by existing diagnostic protocols^7,22^. Genetic correlation analyses indicated that while these factors are indeed genetically correlated with AD, less than approximately 4% of SNP heritability of AD is explained by either factor, indicating that they are not by themselves sufficient to account for the polygenic component of AD genetic architecture. Longitudinal genomic research of neurocognitive change preceding and predicting eventual clinical AD will be of particular value for identifying pathways that may contribute to the polygenic risk for AD pathophysiology.

Our observation that the common variant genetic architecture of AD outside of the *APOE* region is distributed relatively evenly across the genome might be viewed as conflicting with recent findings by Zhang et al.^23^ who, using a Bayesian mixture model (SBayesR), concluded that “the number of causal common SNPs for [AD] may be less than 100, suggesting LOAD is more oligogenic than polygenic.” While the current study makes no specific prediction with respect to the number of causal common SNPs for AD, we do note that the SBayesR approximates the distribution of SNP effects via a mixture of a point mass distribution of null SNPs and three normal distributions of non-null SNP effects. It is possible that, at current sample sizes, it is difficult to distinguish a distribution of variants of very small effect from a point mass distribution of variants with null effect within the SBayesR framework; Given that the primary function of SBayesR is to maximize polygenic prediction, this distinction may be immaterial in that context. Further increases in sample size may be needed to detect the presence of additional causal loci of small effect, and to extend investigations of the genetic architecture of AD into the rare allele spectrum.

It is important to consider that the estimates of common variant SNP heritability from our multivariate modeling are based on summary-based methods that rely on integration of summary data across many different cohorts. Between-cohort variability in the genetic architecture of AD across cohorts, and in methods for ascertainment and diagnosis can lead to attenuation of heritability during the aggregation process. Raw data based estimates of heritability using more homogeneous, yet smaller, cohorts^23^ have often produced even larger heritability estimates than those reported here. This discrepancy could also be explained by the distribution of SNP effects of AD. If the core pathways for AD consist of a small number of causal variants with relatively large effects, then summary-based approaches that rely on assumptions of high polygenicity and normally distributed SNP effects may be yield underestimates of SNP heritability.

Finally, our estimates for the range of plausible values for the heritability of biological AD are based on assumptions regarding the extent to which the genetic signal represented in individuals diagnosed with clinical AD can be extrapolated to individuals with latent biological AD. The most valid insights into the genetic architecture of biological AD will be gleaned from the large scale genomic analysis of direct measures of AD pathophysiology using, for example, positron emission tomography^18^.

## Method

### Bias in Conventional Meta Analyses Combining GWAS and GWAX

Recent large scale meta-analyses combining direct-GWAS and GWAX data have used either inverse variance weighted meta-analysis of regression coefficients^3,24^ or sample size weighted meta-analysis of Z statistics^4,14^. For the inverse variance weighted approach, it is well-known that because of the indirect nature of the GWAX, the regression coefficients and associated standard errors (the squares of which represent the sampling variances used for inverse variance weighting) must be multiplied by a correction factor prior to meta-analysis^1^. The standard correction factor for GWAX of the phenotype of a single first degree relative using the offspring genotype is 2 to correct attenuation due to 50% genetically relatedness. Although not commonly implemented in the literature, we show in S3 of the Supplementary Note that a correction must also be made when implementing the Z statistic approach. We show that for a continuous trait, the sample size must be divided by the square of the correction factor (e.g. by 4 for the standard correction) when implementing the Z statistic approach, with further corrections to sample size needed for case-control traits in ascertained samples.

Naive analysis of GWAX summary statistics produces SNP heritability estimates that are downwardly biased by 75% or more. This bias will carry forward in meta-analysis combining direct GWAS with GWAX statistics (see S4 of the Supplementary Note and simulation below). This will occur when estimating SNP heritability from summary statistics even when regression coefficients and SEs have been corrected, because methods for estimating SNP heritability typically rely on the ratio of the regression coefficients to their standard errors (i.e. the Z statistics), which is preserved under the correction. We show that entering the effective sample size (obtained by dividing the observed N by the square of the correction factor) will produce the unbiased SNP heritability estimate when the appropriate correction factor is known (if the phenotype is a case-control trait, a conversion of the heritability estimate from the observed scale to the liability scale is still called for).

Whether the standard correction factor of 2.0 for GWAS regression coefficients is appropriate, and if not, how the appropriate correction factor can be identified is an unaddressed topic. The standard correction factor is derived simply on the basis of the 50% attenuation to regression estimates expected when proxy cases are first degree relatives of the genotyped individuals. However, attenuation will be more severe, such that the standard correction is insufficient, under a wide range of circumstances, such as those in which: a proportion of genotyped individuals report on the phenotypes of their step or adoptive parents (such that average genetic relatedness of phenotyped and genotyped individuals falls below 50%), when individuals are not well-informed about, misremember or confuse their parents’ phenotype or disease status (such that heritability of the GWX phenotype is attenuated, or contaminated by other heritable phenotypes), or when the average quality of the diagnostics is of lower quality for parental history reports than for direct GWAS of carefully screened case-control sample (such that heritability of the GWAX phenotype is attenuated, or contaminated by other heritable phenotypes). We derive the expected attenuation bias to both regression coefficients and heritability and genetic covariance estimates analytically in S3 and S4 of the Supplementary Note. Below, we introduce a data-driven multivariate approach for meta-analyzing GWAX and direct GWAS summary data that estimates the amount of attenuation in the GWAX (and thus the appropriate correction) directly from the data. Our approach formally models the genetic covariance structure of the GWAX and direct GWAS summary data in order to produce unbiased estimates of SNP heritability and individual SNP effects without manual correction of effect size estimates, standard errors, or sample sizes.

### A Relaxed Multivariate Model for Combining GWAS with GWAX

We use Genomic SEM to estimate a multivariate model of genetic risk for AD using summary data from three sources: direct GWAS, maternal GWAX, and paternal GWAX. In our multivariate model, the total genetic propensity toward AD risk is represented as a latent factor, *F*, that is specified to affect the direct GWAS phenotype and two GWAX phenotypes according to the following system of regression equations

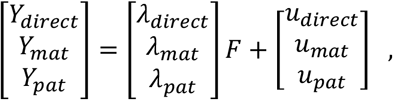

or more compactly as

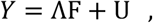

where Λ is a vector of coefficients relating *F* to measured phenotypes *Y*, and U constitutes residual genetic propensities toward each of the measured phenotypes that are independent of *F*, and uncorrelated with one another and with *F*. This model is represented as a path diagram in Supplementary Figure S1 (excluding the dashed portion).

This model implies that the genetic covariance matrix for Y_*direct*,_ Y_*Mat*,_ and Y_*Pat*_ is given as

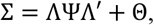

where Ψ represents the covariances among the factors (and in this case contains a single element, representing the variance of F, i.e. 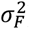) and Θ represents the covariances among the residuals, U (in this case a 3×3 diagonal matrix with diagonal elements 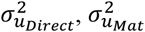, and 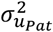).

We specify the model with the minimal identification constraint that *λ*_*direct*_ = 1 such that *F* takes on the scale of the direct GWAS phenotype, and 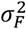 can be interpreted as an unbiased estimate of 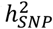 of the meta-analyzed phenotype in the direct GWAS metric. Under this parameterization the departure of *λ*_*Mat*_ and *λ*_*Pat*_ from.5 indicates departure of the empirical data from the standard GWAX model. We also consider an alternative parameterization in which we specify the model with the minimal identification constraint that 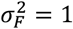 such that the variance of the latent factor F is standardized, and the freely estimated term *λ*_*direct*_ can be interpreted as an unbiased estimate of 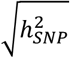 of the meta-analyzed phenotype in the direct GWAS metric. Under this parameterization, departure of *λ*_*Mat*_/*λ*_*direct*_ and *λ*_*Pat*_/*λ*_*direct*_ from.5 indicates departure of the empirical data from the standard GWAX model). Both parameterizations produce equivalent, just identified models, with 0 degrees of freedom, but differ in how parameters must be interpreted. These models can straightforwardly be extended to estimate genetic correlations between *F* and external GWAS phenotypes, such as educational attainment.

#### Estimation of SNP Effects for Multivariate GWAS

The multivariate model can be expanded to include the effect of an individual genetic variant, *x*, on the latent factor, *F* (Supplementary Figure S1, including the dashed portion). Such a model comprises the following two sets of equations:

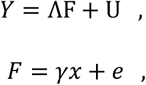

Where *ϒ* is an unstandardized regression coefficient and *e* is a residual. Such a model can be run for all available SNPs, one at a time, such that a complete set of meta-analytic summary statistics for *F* (in this case, AD), can be produced from the relaxed multivariate model. We use the minimal identification constraint *λ*_*direct*_ = 1, such that ϒ takes on the scale of the direct GWAS. To avoid the potential for variability in the optimization of the Λ and Θ parameters that may obscure interpretation of the ϒ coefficients across SNPs, we fixed these paraemters to their values from the model without SNPs when estimating the model for each SNP in our empirical analysis. The resulting summary statistics from this multivariate GWAS serve as an alternative to those produced by more constrained Z statistic-based and (both corrected and uncorrected) inverse variance-based approaches.

#### Model Estimation

Models are estimated in Genomic SEM^15^ using a two-stage approach. In the first stage, the empirical liability-scale genetic covariance matrix S and its sampling covariance matrix V are estimated using a multivariate version of LDSC. When the model to be fit includes individual SNP effects, the S matrix is expanded to include a vector of genetic covariances between the SNP and each of the GWAS and GWAX phenotypes that is derived directly from the univariate GWAS and GWAX estimates. The associated V matrix is also expanded using cross-trait intercepts from LDSC in order to take any potential sample overlap (known or unknown) and/or shared stratification implied by the LDSC model into account. In the second stage, the model is fit to the S matrix, and free parameters are estimated such that they minimize the discrepancy between Σ and S using the diagonally weighted least squares (WLS) fit function with sandwich correction, with weights derived from V, as described in Grotzinger et al.^15^

Note that the liability scale heritability is sensitive to assumptions about the population prevalence of AD. In our analyses of empirical data, our primary models set the population prevalence rate of AD to 4.31% used by Jansen et al.^14^ (corresponding to the population prevalence of Clinical AD at approximately age 70-75 years, the approximate mean age of onset for cases in the direct GWAS of AD), but we consider sensitivity of liability scale heritability estimates to other values, and to the possibility of undiagnosed AD within control participants (see Prevalence Rates in Relation to the Heritability of Clinical AD and AD Pathology, below). We also specifically compare estimates to those obtained with a population prevalence of 5% used by Wightman et al.^4^ to produce their estimate of h^2^ _SNP_ of 2.5%.

### Simulation Study of SNP Heritability

#### Simulation of Summary Statistics

We simulated genome-wide summary statistics for direct GWAS and maternal and paternal GWAX, independently manipulating 1) the sample size for direct GWAS and maternal and paternal GWAX summary statistics, and 2) the relative effect sizes for maternal and paternal GWAX compared to the direct GWAS (i.e. the attenuation coefficients, *λ*). We additionally manipulated heritability of the trait 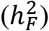 for some conditions to reflect the values estimated in our analyses of empirical data. Table S1 provides the details of the experimental design of our simulation study. We simulated data for three phenotypes (direct GWAS, maternal GWAX, and paternal GWAX) per replication, and 100 replications per each of conditions, for a total of 2700 simulated sets of summary statistics. (As described below, each set of three summary statistics is then analyzed according to three meta-analytic methods, for a total of 2700 meta-analyses). We used the parameter specifications provided Table 1 to produce implied population-level genetic covariance matrices, Σ, calculated as

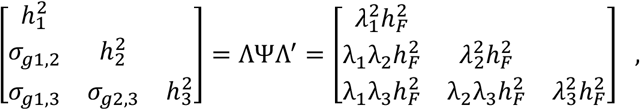

Where 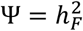 and Λ is the vector of attenuation coefficients (i.e. factor loadings, *λ*). We then used the LD scores for European population for M = 1,184,461 common Hapmap3 SNPs (MAF>.01 excluding the MHC region) provided by Bulik-Sullivan et al., (2015), to simulate summary data for three phenotypes according to the multivariate LDSC equation, i.e.

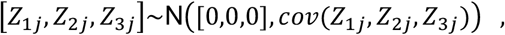

where

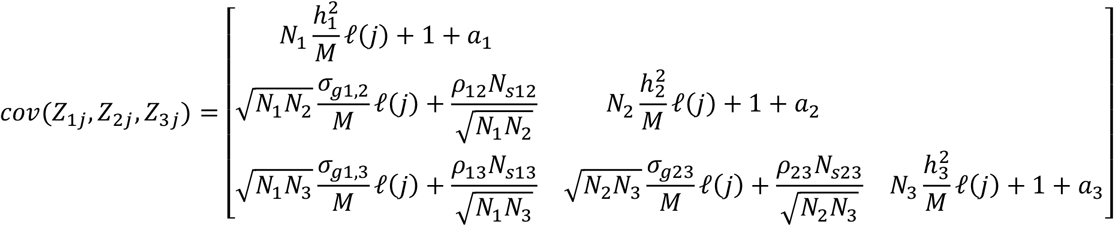

and [*Z*_1*j*,_ *Z*_2*j*,_ *Z*_3*j*_] represents the *Z* statistics for the three GWAS/GWAX phenotypes, *N* is the sample size of the corresponding GWAS/GWAX, *M* is the number of SNPs including in the LD file, ℓ (j) is the LD score of SNP j (that is, the sum of squared correlations between the SNP and all other SNPs), N _s_ is the number of overlapping individuals in the corresponding GWAS/GWAX, *ρ* is the phenotypic correlation within the overlapping individuals, and α represents unmeasured sources of confounding such as population stratification

Our simulation approach, in which summary statistics are directly generated from the LDSC equation is particularly well-suited to our purposes. Direct generation of summary statistics allows us to consider an expansive set of replications and conditions (i.e. 2700 sets of simulated summary statistics total) that would be computationally prohibitive to simulate using a framework in which raw phenotype data were first generated for individual genomes and then submitted to GWAS. Summary data simulated under the LDSC model has the properties needed for analysis of genetic architecture by LDSC and for meta-analysis of effect sizes across phenotypes on a per-variant basis. Importantly, like any simulation approach, our approach also lacks some nuances of real data. For example, while we simulate summary data as a function of LD scores, we do not simulate summary data directly according to linkage disequilibrium (LD) structure (this would require the expansion of the covariance matrix of Z statistics across phenotypes to include cross-SNP covariances, which would dramatically increase computational burden). Thus, the simulated summary data are not appropriate for applications that are directly based on LD structure such as clumping and pruning, identification of lead SNPs within loci, or estimation of heritability using methods such as HDL^25^ and HESS^20^, which directly rely on LD structure (rather than simply on LD scores).

We present results of simulations in which we generate summary statistics under conditions of no sample overlap (*N*_*s*_ = 0) and no population stratification (*a* = 0). However, within this analytic framework, and in previous work using raw data simulation^15^ we have confirmed that Genomic SEM produces unbiased estimates and standard errors when summary statistics are generated with sample overlap, phenotypic correlation within overlapping individuals, and population stratification.

#### Analysis of Simulated Summary Statistics

We compared the performance of three approaches to recover common variant SNP heritability (h^2^): 1) LDSC of summary statistics from a conventional meta-analysis of Z statistics using the observed sample sizes (“Conventional [Uncorrected]”), 2) LDSC of summary statistics from a conventional meta-analysis of Z statistics using the corrected sample sizes, in which the GWAX sample sizes are entered as one quarter their observed value (“Conventional [Standard Correction]”), and 3) our multivariate Genomic SEM-based approach, using the minimal identification constraint *λ*_*direct*_ = 1 such that 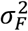 corresponds to the estimate of 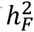). In section S3 of the Supplementary Note, we show that for continuous traits, the uncorrected Z statistic approach is equivalent to the uncorrected inverse variance weighted approach, and the sample-size corrected Z statistic approach (in which the GWAX sample sizes are entered as one quarter their observed values) is equivalent to a corrected inverse variance weighted approach (in which the GWAX beta and its SE are each doubled). We therefore report results for uncorrected and corrected approaches, not distinguishing between Z vs. inverse variance. However, we do note that these approaches are not equivalent for case-control traits when the proportion of cases varies across contributing summary statistics (see sections S2 and S3 of the Supplementary Note). The conventional uncorrected approach is directly reflective of the approaches taken by Jansen et al.^14^ and Wightman et al.^4^.

The conventional corrected approach is directly reflective of the approaches taken by Marioni et al.^3^ and Bellenguez et al.^5^.

For each condition, we calculated: 1) mean parameter estimate, 2) mean standard deviation (SD) of the parameter estimate, 3) mean standard error (SE) of the parameter estimate, and 4) Percent Bias Error (%BE) of the parameter estimate, calculated as:

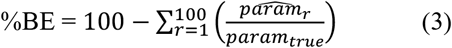

Where 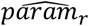 is the parameter estimate in replication *r*, and *param*_*true*_ is its true value for that particular condition.

### Simulation Study of Individual SNP Effects

We extended our simulations for recovery of heritability to allow us to test for the recovery of individual SNP effects. Although the simulation for recovery of heritability generated individual level SNP effects, those analyses simulated based on a population model with a known genetic covariance structure, but not known effects for individual SNPs. However, in order to benchmark performance of each method for estimating individual SNP effects we must know their true effect size in the population from which we draw our simulation. We therefore extended the simulation analyses to include a single individual SNP per replication with a known true effect according to the conditions in Table S1. We set the true effect (ϒ) of this SNP on the F to be equal to the partially standardized (i.e. standard deviations in liability for AD per effect allele) beta coefficient of genome-wide-significant SNP rs60738304 from the IGAP summary statistics (b = 068 MAF = 0.305), such that

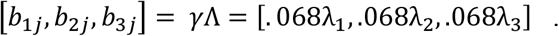

We sampled observed regression coefficients from their sample-size dependent sampling distribution as follows

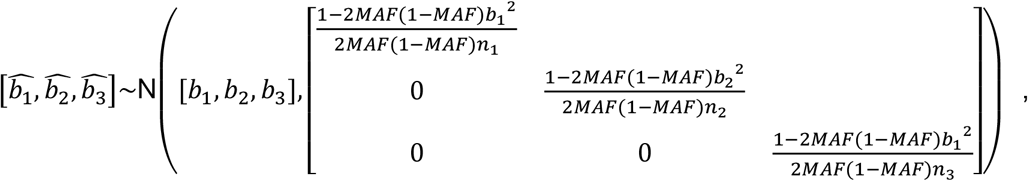

Note that because we simulate under conditions of no sample overlap across any of the direct GWAS and two GWAX datasets, the sampling covariance matrix of the coefficients is diagonal in the population model (the corresponding cells in the V matrix are nevertheless freely estimated when the data are analyzed in Genomic SEM, such that any sample overlap that does exist is automatically detected and corrected for).

#### Analysis of Simulated Data

We computed meta-analytic estimates of *γ* using the corrected and uncorrected approaches, and the multivariate Genomic SEM approach (model with individual SNP effects using the minimal identification constraint *λ*_*direct*_ = 1, such that *γ* takes on the scale of the direct GWAS). Because the Genomic SEM approach requires a genetic covariance matrix that includes terms for individual SNP effects and terms for genome-wide genetic covariances (the genome-wide covariance structure informs the estimation of individual SNP effects), we converted the simulated regression coefficients to SNP-level genetic covariances and appended them to genome-wide genetic covariance matrices estimated from using the genome-wide summary statistics produced under the simulation stud of SNP heritability for the corresponding condition.

For each simulation condition, we calculated: 1) mean parameter estimate, 2) mean standard deviation (SD) of the parameter estimate, 3) mean standard error (SE) of the parameter estimate, 4) mean Z statistic of the parameter estimate, and 5) Percent Bias Error (%BE) of the parameter estimate.

### Selection of GWAS and GWAX Summary Data

We compiled summary data from three published European ancestry direct case-control GWAS of AD and GWAX of parental history of AD. Direct case-control GWAS summary statistics encompassed the discovery sample from the IGAP consortium^10^ comprising 21,982 Clinical AD cases (mean age of onset = 72.93 years) and 41,944 controls (mean age of evaluation = 72.415 years). Cohorts contributing to IGAP varied considerably in the extent to which clinical determination of case and control status was confirmed by autopsy. GWAX summary data of proxy-phenotype AD included 27,696 cases and 260,980 controls of history of maternal AD, and 14,338 cases and 245,941 controls of history of paternal AD, both phenotypes from UK Biobank^3^. Case status for UK Biobank was determined by response to the following two questions “Has/did your father ever suffer from Alzheimer’s disease/dementia?” and “Has/did your mother ever suffer from Alzheimer’s disease/dementia?” at the initial assessment visit (2006–2010), the first repeat assessment visit (2012–2013) and the imaging visit (2014+). Participants whose parents were younger than 60 years or died prior to age 60 years, and without parental age information were excluded. Zhang et al.^23^ report that the mean age of maternal cases was 83.7 years, the mean age of paternal cases was 81.8 years, the mean age of maternal controls was 78.1 years, and the mean age of paternal controls was 76.2 years. Further details on case ascertainment, genotyping, and quality control can be found in the original articles for the corresponding summary statistics.

We additionally curated the following for estimation of genetic correlations: two recent meta-analyses of direct GWAS and GWAX of AD^3,14^, brain volume^26^ in the general population, educational attainment^27^ in the general population and a general genetic factor of cognitive function^28^ in the general population.

### Quality Control of GWAS and GWAX Summary Data

#### LD-Score Regression (LDSC)

All summary statistics used for LDSC were cleaned and processed using the munge function of Genomic SEM, using the standard practice of retaining all HapMap3^29^ SNPs outside of the major histocompatibility complex (MHC) region with minor allele frequencies (MAFs) ≥.01 and information scores (INFO) >.9. To avoid misfit in LD score regression due to extremely large effect sizes within the *APOE* region, we additionally remove this region (CHR19:45,116,911-46,318,605). This has a similar effect to the standard practice in LDSC of removing SNPs with extremely large test statistics. The LD scores used for the analyses presented were estimated from the European sample of 1000 Genomes, but restricted to HapMap3 SNPs as these tend to be well-imputed and produce accurate estimates of heritability. In the LDSC equation, we enter the total number of SNPs in the reference LD panel before excluding the MHC and *APOE* regions, such that the SNP heritability estimates reported here retain similar interpretation to those typically produced by LDSC in other contexts. Based on these procedures, the SNP heritability estimates produced by LDSC should be considered estimates of heritability explained by common SNPs, not including the extremely strong effect of *APOE*.

#### Multivariate GWAS

For the SNP portion of the multivariate model that included individual SNP effects, we used the default QC procedures in Genomic SEM^15^ of removing SNPs with an MAF <.005 in the 1000 Genomes Phase 3 reference panel and SNPs with an INFO score < 0.6 in the univariate GWAS summary statistics. Using these QC procedures left 7,192,577 SNPs across the three contributing summary statistics datasets. Prior to running any multivariate GWAS, all SNP effects were converted to logistic regression coefficients, standardized with respect to the total liability scale variance in the outcome using the sumstats function in GenomicSEM, and corrected for genomic inflation by multiplying the standard errors by square root of the univariate LDSC intercept when the intercept was above 1. These transformed estimates were then multiplied by the SNP variance (estimated as 2 * MAF(1-MAF) to produce genetic covariances. In the case of some variants within the *APOE* region with very large effects, we reduced the MAF relative to its reference panel value when calculating the SNP variance in order to prevent the full S matrix (containing both SNP effects and genetic covariances among the direct GWAS and two GWAX phenotypes) from being nonpositive definite. As we ultimately estimate SNP effects in per effect-allele units, rather than in per standard deviation units, this decision does not bias the estimates of interest.

### Post GWAS Analysis of Meta-Analytic Results in FUMA

We applied FUMA^30^ to results of our multivariate GWAS meta-analysis to identify independent significant SNPs, lead SNPs, and risk loci, using the defaults. Independent significant SNPs were defined as genome wide significant SNPs that are independent from one another at *r*^2^ < .60. Independent significant SNPs are clumped to identify lead SNPs, which are independent from each other at R^2^ < 0.1. In defining genomic risk loci, independent significant SNPs that are associated *r*^2^ ≥ .10 are assigned to the same locus, and independent significant SNPs closer than 250kb are merged into a single locus. The most significant lead SNP in a locus is used to represent that locus. The EUR population from 1000 Genomes Phase 3 was used as the reference panel. As 16 SNPs in the *APOE* block (rs429358) produced very large Z statistics such that their p-value was treated by FUMA as 0 and excluded, we manually entered the most significant SNP in the *APOE* block into FUMA as the lead SNP.

### Prevalence Rates in Relation to the Heritability of Clinical AD and Biological AD

#### Clinical AD

When methods for estimating SNP heritability, such as LDSC, are applied to GWAS of binary phenotypes, the estimate of heritability are on the *observed scale*. Observed scale heritability is difficult to interpret, because it takes on a metric that is idiosyncratic to the balance of cases and controls in the sample, and agnostic to the population prevalence rates of the categories. It is more interpretable to convert observed scale heritability to liability scale heritability based on a liability threshold model, which assumes that a normally distributed liability toward the binary phenotype exists in a population. Under this model, individuals with a liability above a certain threshold are affected whereas those below this threshold are unaffected. The liability-scale SNP heritability estimate is therefore an estimate of the proportion of variation in this continuous liability that is explained by all (directly and indirectly tagged) SNPs on which the GWAS is based. For ascertained samples, in which the proportion of cases does not represent their prevalence in the population, and assuming that the control participants are successfully screened, such that they are unaffected by the disorder (an issue that we return to next), the expected liability scale heritability (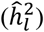) can be obtained from observed scale heritability (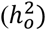)according to Peyrot et al.^31^

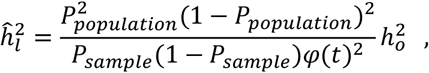

where *P*_*sample*_ is the proportion of cases in the GWAS sample, *P*_*Population*_ is the proportion of cases in the population (i.e. the population prevalence), *t* is the threshold of the cumulative normal distribution corresponding to the population prevalence, and *φ(t)* is the density of the normal distribution at *t*.

Whereas *P*_*sample*_ is known, *P*_*Population*_ must be obtained from collateral epidemiological data. As Yang et al.^32^ write, **“**Application of [the transformation from observed scale heritability to liability scale heritability] requires an estimate of [the population prevalence of the disorder], for which estimates can be surprisingly hard to find, and most applications include a sensitivity analysis to the choice of [this parameter].” Following this guidance, we provide estimates of liability scale heritability across a range of different prevalence rates, and provide rough age equivalents of these prevalence rates based on published epidemiological data from the Medical Research Council Cognitive Function and Ageing Study II^16^. We refer to this estimate as the estimate of heritability of Clinical AD, in that this is the estimate based on the prevalence rate of the clinical diagnosis, and does not account for undetected biological AD that may be more prevalent in the population.

#### Biological AD

The past decade of research on AD has been increasingly cognizant of the fact that “the pathophysiological process of [AD] begins years, if not decades, before the diagnosis of dementia”^33^. Those presenting with Clinical AD may not constitute those individuals with the most severe forms of AD pathophysiology, but may be constitute a group whose cognitive impairments are more overt or detectable due to ancillary characteristics (such as those related to peak premorbid levels of cognitive functioning) that are unrelated to the genetics of biological AD^22^. Indeed, this common observation that differences in ancillary factors such as educational attainment are related to differences in Clinical AD rates among individuals with equivalent levels of brain pathology has led to the popular concept of *cognitive reserve*^7,19,34^. In order to estimate the heritability of biological AD, we capitalize on the Clinical AD GWAS and the age-specific rates of biological AD. We employ the formula for converting observed to liability scale heritability that corrects for case contamination in control participants provided by Peyrot et al.^31^, further adapting to allow for imperfect screening of participants (i.e. screening for Clinical but not biological AD):

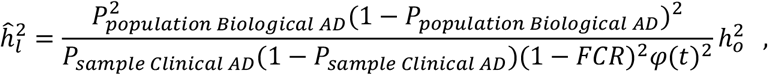

where *FCR* is the false classification rate: the proportion of incorrectly classified control participants in the GWAS. When control participants are a random sample of the population, we set *FCR* = *P*_*Population*_ However, because controls have been screened for clinical AD but not biological AD, and we estimate the liability scale heritability for biological AD under the assumption that clinical AD represents a random subset of biological AD, and compute *FCR* as

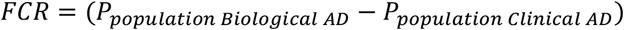

We provide estimates of liability scale heritability of biological AD according to this equation across a range of different population prevalence rates for biological AD, and provide rough age equivalents of these prevalence rates based on recently published data from Jack et al.^18^. To achieve this, we fix *P*_*Population clinical AD*_ the approximate age-specific estimate of the clinical AD prevalence rate from epidemiological data for the mean age of the control sample in IGAP (72.42 years, prevalence rate = 0.0285) to reflect the expected proportion of participants that have been screened out of the control sample for clinical AD) and we vary *P*_*Population biological AD*_

### Local SNP Heritability in HESS

We used HESS to estimate local common variant SNP heritability in 1703 approximately independent blocks across the genome, using common variants (MAF>.01) in the EUR population from 1000 Genomes as the reference panel^20^. Because LDSC relies on the variation in average LD across SNPs to estimate SNP heritability, it cannot be used to estimate local heritability within individual loci in which LD is relatively homogeneously high. In contrast, because HESS relies on the LD matrix itself to estimate SNP heritability, and does not specifically rely on variation in average LD, it is more appropriate for estimating local heritability.

## Supporting information

Supplementary Results

## Data Availability

The data that support the findings of this study are all publicly available or can be requested for access. Specific download links for various datasets are directly below.
Summary statistics for IGAP are available from:
https://www.niagads.org/datasets/ng00075
Summary statistics for UK Biobank can be downloaded from:
https://www.ccace.ed.ac.uk/node/335

https://www.niagads.org/datasets/ng00075

https://www.ccace.ed.ac.uk/node/335

## Acknowledgements

This work presented here would not have been possible without the enormous efforts put forth by the investigators and participants from the International Genomics of Alzheimer’s Project (IGAP) and the UK Biobank. The work from these contributing groups was supported by numerous grants from governmental and charitable bodies. Research reported in this publication was supported by the National Institutes of Health (NIH) grant R01AG054628. J.F. and E.M.T-D. are members of the Population Research Center (PRC) and Center on Aging and Population Sciences (CAPS) at The University of Texas at Austin, which are supported by National Institutes of Health (NIH) grants P2CHD042849 and P30AG066614, respectively. R.E.M. was supported by Alzheimer’s Research UK major project grant ARUK-PG2017B−10.

## Author contributions

J.F. & E.M.T.-D. conceived of the idea, designed the study and formulated the analytic plan. J.F. performed the analyses, with contributions from A.D.G. and E.M.T.-D. J.F. & E.M.T.-D. wrote the paper. All authors contributed critical feedback and contributed to editing the paper.

## Code Availability

GenomicSEM software is an R package that is available from GitHub at the following URL: https://github.com/GenomicSEM/GenomicSEM

Directions for installing the GenomicSEM R package can be found at: https://github.com/GenomicSEM/GenomicSEM/wiki

## Data Availability

The data that support the findings of this study are all publicly available or can be requested for access. Specific download links for various datasets are directly below.

Summary statistics for IGAP are available from: https://www.niagads.org/datasets/ng00075

Summary statistics for UK Biobank can be downloaded from: https://www.ccace.ed.ac.uk/node/335

