## Supplementary Note for "Multivariate Modeling of Direct and Proxy GWAS Indicates Substantial Common Variant Heritability of Alzheimer’s Disease"

Javier de la Fuente,

Andrew D. Grotzinger, Riccardo E. Marioni, Michel G. Nivard,

& Elliot M. Tucker-Drob

### S1. Key Statistics

The standard additive GWAS model estimates coefficient b from the linear regression of quantitative phenotype on genetic variant *x*, written as:

$$y = \mathrm{b}x + e \quad .$$

The sampling variance $\sigma_b^2$ (i.e. the squared standard error, *SE*) of the b estimate is given as

$$\sigma_b^2 = (SE_b)^2 = \frac{\sigma_e^2}{\sigma_x^2 n} = \frac{\sigma_y^2 - \sigma_x^2(b^2)}{\sigma_x^2 n} \quad ,$$

where $\sigma_e^2$ is the residual variance of y, *n* is the sample size, $\sigma_y^2$ is the variance of y, and $\sigma_x^2$ is the variance of the variant. Without loss of generality, we assume that *y* and *x* have each been standardized to mean 0, and standard deviation 1. The sampling variance $\sigma_b^2$ of the b estimate therefore reduces to

$$\sigma_b^2 = (SE_b)^2 = \frac{\sigma_e^2}{n} = \frac{1 - b^2}{n} \quad .$$

Because GWAS effect sizes for complex traits are extremely small, the sampling variance can be closely approximated by

$$\sigma_b^2 = (SE_b)^2 \approx \frac{1}{n} \quad .$$

Finally, the Z statistic for b is equal to the ratio of b to its SE, such that

$$Z = \frac{b}{SE} = \frac{b}{1/\sqrt{n}} = \sqrt{n}\, b \quad .$$

### S2. Relation between Inverse Variance Weighted Meta-Analysis and Meta-Analysis of Z Statistics

Next, we consider two commonly used fixed meta-analysis approaches for combining GWAS data. In this section, we do not consider corrections for sample overlap that have sometimes been implemented, as such corrections are not directly germane to this comparison. (Also note that such corrections are more commonly necessary when meta-analyzing data across multiple case-control GWAS samples, as opposed to meta-analyzing GWAS with GWAX data, which is the focus here.)

*Inverse Variance Approach*

The inverse variance weighted approach can be considered an explicit method, in that it directly combines effect size estimates, b, across *k* sets of GWAX and/or GWAS summary statistics. This approach can be written as the weighted average of regression coefficients, with weights $w_k = \frac{1}{\sigma_{b_k}^2} = n_k$, such that

$$b_{inv\,var} = \frac{\sum w_k b_k}{\sum w_k} = \frac{\sum \frac{1}{\sigma_{b_k}^2} b_k}{\sum \frac{1}{\sigma_{b_k}^2}} = \frac{\sum n_k b_k}{\sum n_k} \;,$$

where $b_{inv\,var}$ is the meta-analytic estimate of b, and $\sum(\;)$ refers to the summation of the relevant terms across each set of summary statistics. Because this approach weighs estimates by the inverse of their squared standard error, it provides greater weight to more precise estimates.

The standard error of the inverse variance weighted meta-analytic estimate is given as

$$SE_{b_{inv\,var}} = \frac{1}{\sqrt{\sum w_k}} = \frac{1}{\sqrt{\sum \frac{1}{\sigma_{b_k}^2}}} = \frac{1}{\sqrt{\sum \frac{1}{1/n_k}}} = \frac{1}{\sqrt{\sum n_k}} \;,$$

such that the Z statistic is

$$Z_{inv\,var} = \frac{b_{inv\,var}}{SE_{b_{inv\,var}}} = \frac{\frac{\sum n_k b_k}{\sum n_k}}{\frac{1}{\sqrt{\sum n_k}}} = \frac{(\sqrt{\sum n_k}\,)(\sum n_k b_k)}{\sum n_k} = \frac{\sum n_k b_k}{\sqrt{\sum n_k}}$$

*Z Statistic Approach*

A second common approach for meta-analysis of GWAX and/or GWAS summary statistics uses Z statistics and sample sizes at the input. (This approach also accommodates *p* values, so long as the direction of effect is known, as such information can be used to back out the relevant Z statistics).

The Z statistic approach can be considered an indirect method, in that although Z and *n* are taken as the input, this approach is mathematically equivalent to an inverse variance weighed meta-analysis of standardized linear regression estimates across datasets. This approach is popularly implemented in METAL software[1], and serves as the basis for the multivariate GWAS meta-analysis of AD introduced in Jansen et al.[2] and used in the most recent AD GWAS-GWAX meta-analysis by Wightman et al.[3]. It can be written as the weighted average of Z statistics, with weights $w_k = \sqrt{n_k}$.

$$Z_{Z\ Stat} = \frac{\sum w_k\, Z_k}{\sqrt{\sum w_k^2}} = \frac{\sum \sqrt{n_k}\, Z_k}{\sqrt{\sum n_k}} \quad .$$

Recalling that $Z = \sqrt{n}\; b$ , we therefore have

$$Z_{Z\ Stat} = \frac{\sum \sqrt{n_k}\, Z_k}{\sqrt{\sum n_k}} = \frac{\sum \sqrt{n_k}\, (\sqrt{n_k} b_k)}{\sqrt{\sum n_k}} = \frac{\sum n_k b_k}{\sqrt{\sum n_k}} \quad ,$$

which is equivalent to the Z statistic for the inverse variance weighted meta-analysis ($Z_{inv\ var}$) that is given above. Thus, the Z statistic approach to meta-analysis is equivalent to an inverse variance weighted meta-analysis of standardized regression coefficients. This equivalence can further be seen as follows

$$Z_{Z\ Stat} = \frac{\sum n_k b_k}{\sqrt{\sum n_k}} = \frac{\sqrt{\sum n_k}}{\sqrt{\sum n_k}} \frac{\sum n_k b_k}{\sqrt{\sum n_k}} = \left(\sqrt{\sum n_k}\right) \frac{(\sum n_k b_k)}{\sum n_k}$$

$$= \frac{\frac{\sum n_k b_k}{\sum n_k}}{\frac{1}{\sqrt{\sum n_k}}} = \frac{\frac{\sum \frac{1}{\sigma_{b_k}^2} b_k}{\sum \frac{1}{\sigma_{b_k}^2}}}{\frac{1}{\sqrt{\sum \frac{1}{\sigma_{b_k}^2}}}} = \frac{b_{inv\ var}}{SE_{b_{inv\ var}}} \quad .$$

*Binary Phenotypes*

The above derivations are developed specifically for linear regression, and thus are most relevant to the analysis of continuously distributed quantitative phenotypes. When the phenotype under investigation is a binary "all-or-nothing" phenotype, taking on one of two values (e.g. 0 or 1), such as a case-control phenotype, a logistic regression is more appropriate for relating the effect of the variant on the unobserved continuous liability for the binary outcome. However, in the context of GWAS of complex traits where individual SNP effects are extremely small, the regression coefficient for the logistic regression of the binary phenotype on the standardized variant ($b_{logit\ STDX}$) can be closely approximated from the coefficient from the linear regression of the standardized binary phenotype on the standardized variant X ($b_{linear\ STD}$) as follows[4,5]

$$b_{logit\ STDX} \approx \frac{b_{linear\ STD}}{\sqrt{v(1-v)}} \ ,$$

$$\sigma^2_{b_{logit\ STDX}} \approx \frac{\sigma^2_{b_{linear\ STD}}}{v(1-v)} = \frac{1}{v(1-v)n} \ ,$$

where $v$ is the proportion of cases, such that $v(1\text{-}v)$ is the observed variance of the binary phenotype. Note that in this section we use notation to make explicit whether a regression coefficient represents a linear or logistic coefficient, and whether it is fully standardized (STD) or only standardized with respect to the genetic predictor (STDX), as these distinctions are critical to proper interpretation and application of the formulae presented.

The fully standardized coefficient ($b_{linear\ STD}$) for the linear regression of a binary trait on a variant, is sometimes referred to as representing the "observed scale," and is the basis for the concept of observed scale heritability. Thus, a key interpretational consideration when using the Z statistic approach to meta-analyze binary phenotypes, is that this approach is equivalent to an inverse variance weighted meta-analysis of standardized linear regression coefficients, where coefficients are standardized with respect to the *observed* variance of the binary phenotype, i.e. $v(1\text{-}v)$, where $v$ is the proportion of cases in the corresponding GWAS, rather than the variance of the continuous liability for binary phenotype. However, using the approximations given above, we can specify the Z Statistic approach in terms of logistic regression coefficients instead of standardized linear regression coefficients, as

$$Z_{Z\ Stat} = \frac{\dfrac{\sum \frac{1}{\sigma^2_{b_{linear\ STD_k}}} b_{linear\ STD_k}}{\sum \frac{1}{\sigma^2_{b_{linear\ STD_k}}}}}{\dfrac{1}{\sqrt{\sum \frac{1}{\sigma^2_{b_{linear\ STD_k}}}}}} = \frac{\dfrac{\sum \frac{1}{v_k(1-v_k)\sigma^2_{b_{logit\ STDX_k}}} \sqrt{v_k(1-v_k)} b_{logit\ STDX_k}}{\sum \frac{1}{v_k(1-v_k)\sigma^2_{b_{logit\ STDX_k}}}}}{\dfrac{1}{\sqrt{\sum \frac{1}{v_k(1-v_k)\sigma^2_{b_{logit\ STDX_k}}}}}}$$

$$= \frac{\frac{\sum w_k\left(\sqrt{v_k(1-v_k)}b_{logit\ STDX_k}\right)}{\sum w_k}}{\frac{1}{\sqrt{\sum w_k}}} \ ,$$

where $w_k = \left(v_k(1-v_k)\sigma^2_{b_{logit\ STDX_k}}\right)^{-1}$. Thus, the Z statistic approach will approximate a meta-analysis of a study-specific transformation of the logistic regression coefficient (specifically, the logistic coefficient multiplied by the standard deviation of the binary phenotype) that is weighted both by the inverse sampling variance of the logistic regression coefficients and the inverse variance of the binary phenotype. It may therefore be particularly difficult to interpret the meta-analytic estimate from the Z Statistic approach when the GWAS summary statistics being meta-analyzed differ considerably in the proportion of cases, as will likely be the case for ascertained studies, as the transformation of the logistic regression coefficient will differ for each set of summary statistics. In other words, when case proportions differ across sets of summary statistics, the Z Statistic approach amounts to a meta-analysis of arbitrary study-specific transformations of the effect sizes of interest.

In contrast, when applied to logistic regression coefficients (or to linear regression coefficients that have already been transformed into logistic regression coefficients) the inverse variance weighted approach to meta-analysis of direct GWAS summary data can be straightforwardly interpreted as a direct meta-analysis of the effect sizes of interest. Using the approximations given above, we can specify this approach for logistic regression coefficients that have been standardized with respect to the genetic variant X as follows

$$b_{logit\ STDX_{inv\ var}} = \frac{\sum w_k b_{logit\ STDX_k}}{\sum w_k} = \frac{\sum \frac{1}{\sigma^2_{b_{logit\ STDX_k}}} b_{logit\ STDX_k}}{\sum \frac{1}{\sigma^2_{b_{logit\ STDX_k}}}} = \frac{\sum v_k(1-v_k)n_k b_{logit\ STDX_k}}{\sum v_k(1-v_k)n_k} \ ,$$

$$SE_{b_{logit\ STDX\ inv\ var}} = \frac{1}{\sqrt{\sum w_k}} = \frac{1}{\sqrt{\sum v_k(1-v_k)n_k}} \ .$$

Next, we elucidate how further considerations must be given when, rather than only consisting of direct GWAS estimates, the summary statistics being meta-analyzed also consist of GWAX estimates. We focus on the meta-analysis of GWAS and GWAX of quantitative phenotypes, but we note that when the GWAS/GWAX phenotypes are binary, the properties given here additionally apply. Note that we address issues surrounding the estimation of heritability of binary phenotypes in section S8.

**S3. Meta-Analyses Combining GWAX and Direct GWAS Data**

We can write the direct GWAS and proxy GWAS regression equations as

$$y_{direct} = b_{GWAS}\, x + e_{direct} \quad ,$$

$$y_{mat} = b_{mat\ GWAX}\, x + e_{y_{mat}} \quad ,$$

$$y_{pat} = b_{pat\ GWAX}\, x + e_{y_{pat}} \quad ,$$

where $x_i$ is genetic variant; $y_{direct}, y_{mat}$ and $y_{pat}$ are the genotyped individual's own phenotype, the maternal phenotype, and the paternal phenotype, respectively; and $e_{direct,i}, e_{mat,i}$ and $e_{pat,i}$ are residuals. Without loss of generality, we assume that the *ys* and *xs* have each been standardized to mean 0, and standard deviation 1, and we assume that the three sets of GWAS/GWAX summary statistics are independent (i.e. the data have not been obtained from trios, and there has not been assortment related to the maternal and paternal AD phenotypes).

Consider a data generating model in which the GWAX regression estimate is used to approximate the estimate $b^*_{GWAS}$ that would have been obtained under a direct GWAS using the parent's own genotypes,

$$b_{mat\ GWAX} = \lambda_{mat} \cdot b^*_{mat\ GWAS} \quad ,$$

$$b_{pat\ GWAX} = \lambda_{pat} \cdot b^*_{pat\ GWAS} \quad ,$$

where the $\lambda s$ represent attenuation coefficient that reduces the expected effect size due to the indirect nature of the GWAX. The standard GWAX model[6] treats $\lambda_{mat} = \lambda_{pat} = .5$, reflecting the assumptions that offspring are 50% related to the parents on whom they report the phenotype (e.g. disease history) and that the phenotype of interest has been measured with equal fidelity in family history report as would be obtained in direct case-control GWAS. These assumptions will be violated when a proportion of genotyped individuals report on the phenotypes of their step or adoptive parents, when individuals are not well-informed about their parents' phenotype or disease status, when individuals misremember or forget, their parent's diagnoses, when individuals confuse their parent's phenotypes or diagnoses (e.g. confusing delirium for AD) , and/or when the quality of the diagnostics is lower for parent history reports than for direct GWAS of carefully screened case-control samples.

The standard GWAX approach is to multiply naïve GWAX coefficients by 2, in order to produce an estimate of the regression coefficient that is on the same scale as that which would have been obtained under a direct GWAS. In order to preserve the *p* value of the estimate, the SE is also multiplied by 2 (the correction of the naïve coefficients, after all, cannot produce an increase in power unless the original SEs of the naïve coefficients were incorrect). It can be seen that when $\lambda_k$ =.5, this correction is appropriate:

$$\hat{b}^*_{GWAS} \quad = 2 \cdot b_{GWAX} = 2 \cdot \lambda \cdot b^*_{GWAS} = \; 2 \cdot .5 \cdot b^*_{GWAS} = \; b^*_{GWAS} \quad .$$

However, it follows that when $\lambda_k$ is less than .5, then the $\hat{b}^*_{GWAS}$ estimate using this correction will be downwardly biased by a factor of $2(.5 - \lambda_k)$.

Consider the situation in which the naïve uncorrected version of the GWAX coefficients are entered into the inverse weighted variance approach. Assuming the standard GWAX assumption that $\lambda_{mat} = \lambda_{pat} = \; .5$, it is clear that this will produce a downwardly biased estimate of b, by virtue of meta-analyzing the effect size of interest from the direct GWAS with effect sizes that are downwardly biased by 50%. This is illustrated as follows

$$b_{inv\,var} = \; \frac{\sum w_k b_k}{\sum w_k} = \frac{w_{direct\,GWAS}\, b_{direct\,GWAS} + w_{mat\,GWAX}\, b_{mat\,GWAX} + w_{pat\,GWAX}\, b_{pat\,GWAX}}{w_{direct\,GWAS} \; + w_{mat\,GWAX} \; + w_{pat\,GWAX}}$$

$$= \frac{w_{direct\,GWAS}\, b_{direct\,GWAS} + w_{mat\,GWAX} \cdot \lambda_{mat} \cdot \; b^*_{mat\,GWAS} \; + w_{pat\,GWAX} \cdot \lambda_{pat} \cdot \; b^*_{pat\,GWAS}}{w_{direct\,GWAS} \; + w_{mat\,GWAX} \; + w_{pat\,GWAX}}$$

$$= \frac{w_{direct\,GWAS}\, b_{direct\,GWAS} + w_{mat\,GWAX}\, (.5)\; b^*_{mat\,GWAS} \; + w_{pat\,GWAX}\, (.5)\; b^*_{pat\,GWAS}}{w_{direct\,GWAS} \; + w_{mat\,GWAX} \; + w_{pat\,GWAX}},$$

where $w_k = \frac{1}{\sigma^2_{b_k}} = n_k$. In other words, when used with naïve GWAX coefficients, the inverse variance weighted approach will produce meta-analytic estimates that are the sample-size weighed average of the unbiased direct GWAS and the biased GWAX. For instance, when the sample sizes for the direct GWAS, maternal GWAX, and paternal GWAX are equal, and the naïve uncorrected version of the GWAX coefficients are entered, $b_{inv\,var}$ is expected to be two thirds (i.e. 66.6%) the true value, under the standard GWAX assumption that $\lambda_{mat} = \lambda_{pat} = \; .5$. Moreover, as $R^2 = b^2$, in this example $R^2$ is expected to be four ninths (i.e. 44.4%) the unbiased $R^2$ value.

When the corrected GWAX coefficients are entered using the standard correction in which the coefficients and their standard errors are each multiplied by 2, we have

$$b_{inv\,var} = \; \frac{\sum w_k b_k}{\sum w_k} = \frac{w_{direct\,GWAS}\, b_{direct\,GWAS} + w_{mat\,GWAX}\, 2\; b_{mat\,GWAX} + w_{pat\,GWAX}\, 2\; b_{pat\,GWAX}}{w_{direct\,GWAS} \; + w_{mat\,GWAX} \; + w_{pat\,GWAX}}$$

$$= \frac{w_{direct\,GWAS}\, b_{direct\,GWAS} + w_{mat\,GWAX}\, 2 \cdot \lambda_{mat} \cdot b^*_{mat\,GWAS} \; + w_{pat\,GWAX}\, 2 \cdot \lambda_{pat} \cdot b^*_{pat\,GWAS}}{w_{direct\,GWAS} \; + w_{mat\,GWAX} \; + w_{pat\,GWAX}}$$

It can be seen that the inverse variance approach yields unbiased estimates of $b$ and $R^2$, so long as the standard GWAX assumption that $\lambda_{mat} = \lambda_{pat} = .5$ is correct. This can be seen as follows

$$b_{inv\,var} = \frac{w_{direct\,GWAS}\, b_{direct\,GWAS} + w_{mat\,GWAX}\, 2(.5) b^{*}_{mat\,GWAS} + w_{pat\,GWAX}\, 2(.5) b^{*}_{pat\,GWAS}}{w_{direct\,GWAS} + w_{mat\,GWAX} + w_{pat\,GWAX}}$$

$$= \frac{w_{direct\,GWAS}\, b_{direct\,GWAS} + w_{mat\,GWAX}\, b^{*}_{mat\,GWAS} + w_{pat\,GWAX}\, b^{*}_{pat\,GWAS}}{w_{direct\,GWAS} + w_{mat\,GWAX} + w_{pat\,GWAX}} \ .$$

However, it can also be seen that when the GWAX assumption is violated, such that that $\lambda_{mat} < .5$ and/or $\lambda_{pat} < .5$, the meta-analytic $b_{inv\,var}$ estimate using the standard correction will be downwardly biased by virtue of the constituent corrected GWAX estimates being downwardly biased by a factor of $2(.5 - \lambda_k)$.

Now consider how the Z statistic approach operates with respect to the meta-analysis of GWAS with GWAX data. As derived earlier, the Z statistic approach is mathematically equivalent to an inverse variance weighed meta-analysis of standardized linear regression estimates across datasets. Because conversion of linear regression coefficients from GWAX to GWAS metric requires that the identical transformation be performed on the standard errors of the associated regression coefficients, the Z statistics associated with this transformation are unchanged. For example in the context of the standard correction of GWAX estimates,

$$\hat{b}^{*}_{GWAS} = 2 \cdot b_{GWAX}$$

$$SE_{\hat{b}^{*}_{GWAS}} = 2 \cdot SE_{b_{GWAX}}$$

$$Z_{\hat{b}^{*}_{GWAS}} = \frac{\hat{b}^{*}_{GWAS}}{SE_{\hat{b}^{*}_{GWAS}}} = \frac{2 \cdot b_{GWAX}}{2 \cdot SE_{b_{GWAX}}} = \frac{b_{GWAX}}{SE_{b_{GWAX}}} = Z_{b_{GWAX}} \ .$$

Therefore, even when the GWAX effect sizes and standard errors have been transformed, the associated Z statistics remain unchanged, and the Z statistic approach to meta-analyzing GWAX together with GWAS, whether transformed or untransformed, will be equivalent to an inverse variance weighed meta-analysis of ***uncorrected*** standardized linear regression estimates across datasets, and will therefore be biased in its uncorrected form.  This is concerning in that the Z statistic approach is often considered more appealing than the inverse variance weighting approach, as it is commonly assumed that this approach does not require effect sizes to be scaled to the same metric across sets of summary statistics. This may generally be the case when the summary statistics being combined are from direct GWAS of quantitative phenotypes. For instance, the Z statistic approach would allow summary statistics from GWAS of unstandardized phenotypes to be combined with those of GWAS of standardized phenotypes without first requiring the unstandardized estimates to be standardized. However, in order to conduct an unbiased meta-analysis of GWAX together with direct GWAS using the Z statistic approach, the sample size for the GWAX must be divided by the square of the correction factor, such that in the context of the standard correction factor of 2 (corresponding to $\lambda = .5$), the sample size is divided by 4. This can be observed by considering how b in inferred from Z and n.

$$b = \frac{Z}{\sqrt{n}} \quad ,$$

$$\lambda \cdot b = \frac{\lambda \cdot Z}{\sqrt{n}} = \frac{Z}{\frac{1}{\lambda} \cdot \sqrt{n}} = \frac{Z}{\sqrt{\left(\frac{n}{\lambda^2}\right)}} \quad ,$$

$$2b = \frac{2 \cdot Z}{\sqrt{n}} = \frac{Z}{.5 \cdot \sqrt{n}} = \frac{Z}{\sqrt{.25 \cdot n}} \quad .$$

Note that transforming the naively-weighted Z statistics approach into a meta-analytic regression coefficient using the pooled version of the corrected *n* will be insufficient for fully removing bias of this approach, as the corrected *n* must be used to form the weights for the constituent summary statistics being meta-analyzed.

**S4. SNP Heritability and Genetic Covariance in Relation to GWAS and GWAX**

Consider a population model in which K phenotypes and M SNPs are measured in N individuals, and modeled according to the hypothetical multivariate multiple regression model:

$$\mathbf{Y} = \mathbf{XB} + \mathbf{E}$$

where **Y** is an N× K matrix of standardized scores for person i on phenotype k, X is an N×M matrix of standardized genotypes for person i on SNP j, B is an M×K matrix of true genotype effect sizes for SNP j on phenotype k, and E is an N×K matrix of residuals for person i on phenotype k. Under this framework, the SNP heritability ($h^2_{SNP}$) of phenotype k is defined as the multiple $R^2$ from its multiple regression on the M SNPs using the true values of the vector of coefficients, $b_k$, contained within **B**, and genetic covariance between any pair of phenotypes is defined as the covariance between the expected values of the phenotypes on the basis of both the M genotypes and the corresponding vectors ($\mathbf{b_k}$) of true regression coefficients contained within **B**.

We can straightforwardly observe the effect of using GWAX data to estimate $h^2$ for phenotype *k*. Given the M×M linkage disequilibrium matrix, L, containing the correlations among genetic variants, and the vector of true effect sizes $\mathbf{b_k}$, the multiple $R^2$ for the population model can be expressed as

$$h^2 = R^2_{pop\ GWAS} = \mathbf{b}_\mathbf{k}'\mathbf{L}\mathbf{b}_\mathrm{k} \quad .$$

Given the indirect nature of the GWAX, the vector $\mathbf{b_k}$ of genetic effects on $\mathbf{y_k}$ is reduced by a factor of $\lambda$ such that

$$h^2_{GWAX} = R^2_{pop\ GWAX} = (\lambda\mathbf{b_k})'\mathbf{L}(\lambda\mathbf{b_k}) = \lambda^2(\mathbf{b}_\mathbf{k}'\mathbf{L}\mathbf{b_k}) = \lambda^2 h^2_{SNP} \quad .$$

Thus, SNP heritability, as estimated from GWAX data, is expected to be reduced by a factor of $\lambda^2$, unless a correction is made. For instance, under the standard GWAX assumption that $\lambda = .5$, uncorrected GWAX estimates of SNP heritability are expected to be one quarter the size of the true SNP heritability, such that the naïve estimate of SNP heritability from GWAX, $h^2_{SNP\ GWAX}$, must be quadrupled to produce an unbiased estimate of $h^2_{SNP}$.

As the genetic covariance ($\sigma_g$) between two phenotypes can be expressed of the genetic correlation ($r_g$) expressed in the metric of their heritability's ($h_1^2$ and $h_2^2$), a similar attenuation also applies to genetic covariance. In other words, for phenotypes 1 and 2, the genetic covariance is

$$\sigma_g = r_g\sqrt{h_1^2}\sqrt{h_2^2} \quad ,$$

such that

$$\sigma_{g\ GWAX} = r_g\sqrt{\lambda_1^2 h_1^2}\sqrt{\lambda_2^2 h_2^2} = \lambda_1\lambda_2 r_g\sqrt{h_1^2}\sqrt{h_2^2} = \lambda_1\lambda_2\sigma_g.$$

For instance, under the standard GWAX assumption that $\lambda_1 = \lambda_2 = .5$, uncorrected GWAX estimates of genetic covariance are expected to be one quarter the size of the true genetic covariance.

In practice, heritability and genetic covariance can be estimated from GWAS summary data using LD Score Regression (LDSC), which treats regression coefficients as phenotype-specific random effects, varying over SNPs[7,8]. The values are estimated by regressing the product of Z statistics for the linear regression of phenotypes 1 and 2 on SNP *j* on the LD score of SNP *j* and solving for $\sigma_g$, as follows

$$E[z_{1j}z_{2j}] = \sqrt{N_1N_2}\frac{\sigma_g}{M}\ell(j) + \frac{\rho N_s}{\sqrt{N_1N_2}} + a \quad ,$$

where $N_1$ and $N_2$ are the sample sizes for phenotypes 1 and phenotypes 2, M is the number of SNPs, $\ell(j)$ is the LD score of SNP $j$ (that is, the sum of squared correlations between the SNP and all other SNPs), $N_s$ is the number of individuals included in both GWAS samples, $\rho$ is the phenotypic correlation within the overlapping samples, and $a$ is a term representing unmeasured sources of confounding such as shared population stratification across GWASs. Note that when the $z$ statistics for the same phenotype are double entered into the left-hand side of the above equation, such that $E[z_{1j}z_{2j}]$ becomes $E[z_j^2] = E[\chi_j^2]$, the equation reduces to the univariate S-LDSC model, and $\sigma_g$ becomes an estimate of $h^2_{SNP}$. A particular strength of LDSC is that it produces estimates of $\sigma_g$ and $h^2_{SNP}$ that are robust to biases that would otherwise result from sample overlap and population stratification[7–9].

In the next section we introduce a multivariate method for empirically estimating the λ attenuation coefficients using a combination of GWAX and direct GWAS summary data. Our method, based in Genomic Structural Equation Modelling, simultaneously estimates and incorporates these λ terms while estimating heritability and genetic covariance, and can be used to directly compute λ-corrected meta-analytic summary statistics for individual variants. Our method does not require manual correction of summary statistics for λ, either by multiplying GWAX effect sizes and their standard errors by a correction factor or by correcting the sample size entered into LDSC. However, it may be instructive to consider how such a correction might be made in the context of the GWAX model, we describe how such a manual correction can be made when estimating $\sigma_g$ (and $h^2_{SNP}$ in the case of univariate LDSC). Simply put, because $\sigma_{g\ 1,2}$ is expected to be biased by a factor of $\lambda_1\lambda_2$, we can produce a manually corrected version of LDSC by entering $\lambda_1^2 N_1$ in place of $N_1$ and $\lambda_2^2\, N_2$in place of $N_2$. The slope of the LDSC regression equation becomes

$$\sqrt{\lambda_1^2 N_1 \lambda_2^2\, N_2}\,\frac{\sigma_g}{M} = \lambda_1\lambda_2\sqrt{N_1 N_2}\,\frac{\sigma_g}{M}\ .$$

Thus, under the standard GWAX assumption that $\lambda_1 = \lambda_2 = .5$, when the sample sizes associated GWAX summary data are entered at one quarter their actual value, the $\sigma_g$ term is correctly estimated at four times larger than it would be under the naïve, uncorrected, LDSC model.

#### S5. A Relaxed Multivariate Model for Combining GWAS with GWAX

Consider a data generating model introduced earlier, in which summary data are available from three sources, direct GWAS, maternal GWAX, and paternal GWAX. We describe this model in terms of linear GWAS and GWAX of quantitative phenotypes, but this description straightforwardly generalizes to binary phenotypes using the liability scale estimates of $\sigma_g$ and $h^2_{SNP}$ and logistic regression coefficients (or linear probability model coefficients that have been rescaled to logistic regression coefficients)[4,5,10].

We can write a model in which the total genetic propensity toward the phenotype *F* is specified to affect the direct GWAS phenotype and two GWAX phenotypes according to the following system of regression equations

$$\begin{bmatrix} Y_{direct} \\ Y_{mat} \\ Y_{pat} \end{bmatrix} = \begin{bmatrix} \lambda_{direct} \\ \lambda_{mat} \\ \lambda_{pat} \end{bmatrix} F + \begin{bmatrix} u_{direct} \\ u_{mat} \\ u_{pat} \end{bmatrix} \ ,$$

or more compactly as

$$Y = \Lambda \mathrm{F} + \mathrm{U} \ ,$$

where, Y constitutes the measured phenotypes, Λ is a vector of attenuation coefficients relating the latent genetic propensity toward the phenotype of interest to measured phenotypes, and U constitutes residual genetic propensities toward each of the measured phenotypes that are independent of *F*, and uncorrelated with one another and with *F*.

Using standard linear structural relations (LISREL) notation[11], the covariances among the independent variables in the model can be specified as two covariance matrices. The covariance matrix Ψ represents the covariances among the factors, and in this case contains a single element, representing the variance of F, i.e. $\sigma^2_F$. The covariance matrix Θ represents the covariances among the residuals, U, and in this case is a 3×3 diagonal matrix with diagonal elements $\sigma^2_{u_{Direct}}$, $\sigma^2_{u_{Mat}}$, and $\sigma^2_{u_{Pat}}$ .

Under this model, the expected genetic covariance matrix for $Y_{Direct}$, $Y_{Mat}$, and $Y_{Pat}$ is given as

$$\Sigma = \Lambda \Psi \Lambda' + \Theta.$$

Estimating $h^2_{SNP}$ using the assumptions of the standard GWAX approach is equivalent to assuming that $[\lambda_{direct} \quad \lambda_{mat} \quad \lambda_{pat}] = [1 \quad .5 \quad .5]$ and $[\sigma^2_{u_{direct}} \quad \sigma^2_{u_{mat}} \quad \sigma^2_{u_{pat}}] = [0 \quad 0 \quad 0]$, and estimating $\sigma^2_F$. This can be achieved in the Genomic SEM framework by first estimating the empirical genetic covariance matrix S using a multivariate version of LDSC and subsequently estimating the free model parameters to minimize the discrepancy between Σ and S using a fit function, such as the weighted least squares (WLS) fit function described in Grotzinger et al.[5], which takes into account the full sampling covariance matrix of the elements within S (labelled V). This is an overidentified model, and may thus incur misfit. However, we are able to relax the above assumptions by freely estimating the Λ terms. We can specify the model to estimate all terms, with the minimal identification constraint that $\lambda_{direct} = 1$ such that the factor takes on the scale of the direct GWAS phenotype, and $\sigma_F^2$ can be interpreted as an unbiased estimate of $h_{SNP}^2$ of the meta-analyzed phenotype in the direct GWAS metric, with the departure of $\lambda_{mat}$ and $\lambda_{pat}$ from .5 indicating departure of the empirical data from the standard GWAX model. (Alternatively, we can specify the model with the minimal identification constraint that $\sigma_F^2 = 1$, such that the variance of the latent factor F is standardized, and the freely estimated term $\lambda_{Direct}$ can be interpreted as an unbiased estimate of $\sqrt{h_{SNP}^2}$ of the meta-analyzed phenotype in the direct GWAS metric, and $\lambda_{mat}$ and $\lambda_{pat}$ representing the attenuation coefficients rescaled to the $\sqrt{h_{SNP}^2}$ metric, such that departure of $\lambda_{mat}/\lambda_{direct}$ and $\lambda_{pat}/\lambda_{direct}$ from .5 indicates departure of the empirical data from the standard GWAX model). This is a 0 df model that is just identified. This factor model implies the following equalities with respect to the genetic covariances as a function of the model parameters

$$\sigma_{direct,mat} = \lambda_{direct}\sigma_F^2\lambda_{mat} \quad ,$$

$$\sigma_{direct,pat} = \lambda_{direct}\sigma_F^2\lambda_{pat} \quad ,$$

$$\sigma_{mat,pat} = \lambda_{mat}\sigma_F^2\lambda_{pat} \quad .$$

When we impose the minimal identification constraint $\lambda_{Direct} = 1$ to set the metric of F (such that $\sigma_F^2$ becomes an estimate of $h_{SNP}^2$ in the direct GWAS metric), and solve for the model parameters as a function of the genetic covariances, amounting to a version of Spearman's method of triads[12,13], we obtain

$$\sigma_F^2 = \frac{\sigma_{direct,mat} \cdot \sigma_{direct,pat}}{\sigma_{mat,pat}} \quad ,$$

$$\lambda_{mat} = \frac{\sigma_{mat,pat}}{\sigma_{direct,pat}} \quad ,$$

$$\lambda_{pat} = \frac{\sigma_{mat,pat}}{\sigma_{direct,mat}} \quad .$$

An important observation from the above is that, although the factor F has been specified to take on the metric of the direct GWAS, the estimate of $\sigma_F^2$ is not specifically or exclusively informed by the SNP heritability estimate from the direct GWAS, but rather by the estimates of genetic covariance between all three indicators ($Y_{direct}$, $Y_{mat}$, and $Y_{pat}$). In fact, the heritability of each indicator only directly informs the residual genetic variances, i.e. the SNP heritability of each indicator that is not explained by the common factor F. This can be seen as follows:

$$\sigma_{u_{direct}}^2 = \sigma_{Y_{direct}}^2 - \sigma_F^2 \quad ,$$

$$\sigma^2_{u_{mat}} = \sigma^2_{Y_{mat}} - \lambda^2_{mat} \cdot \sigma^2_F \quad ,$$

$$\sigma^2_{u_{pat}} = \sigma^2_{Y_{pat}} - \lambda^2_{pat} \cdot \sigma^2_F \quad .$$

One somewhat constrained version of the model that may be desirable is to fix the $\sigma^2_{u_{Direct}}$ parameter to 0 a priori, in order to prioritize the direct GWAS as a direct manifestation of the genetic signal of interest. As shown in the Supplement of Grotzinger et al.[5], such a specification closely resembles the MTAG model[14]. Allowing $\sigma^2_{u_{direct}}$to be freely estimated allows for the possibility that the direct GWAS, like the GWAX, may contain ancillary genetic signal that is not shared across the three indicators (we discuss this topic further under *The Heterogeneity Coefficient*, $Q_{SNP}$ below). In our analysis of AD, we freely estimate $\sigma^2_{u_{direct}}$and find it to be ~0.

**S6. Using the Multivariate Model to Generate Variant-Specific Meta-Analytic Estimates**

The multivariate Genomic SEM approach can be used to perform GWAS-GWAX meta-analysis relaxing the conventional assumptions that $[\lambda_{direct} \quad \lambda_{mat} \quad \lambda_{pat}] = [1 \quad .5 \quad .5]$ in order to produce variant-level summary statistics. Such a multivariate GWAS approach is accomplished by freely estimating parameters from a series of models (one per genetic variant) in which the unmeasured variable F, is regressed on the variant. This model is identified by relying the summary data for direct GWAS and maternal and paternal GWAX, which contain estimates from each of the following equations

$$y_{direct} = b_{direct}\, x + e_{direct} \quad ,$$

$$y_{mat} = b_{mat}\, x + e_{y_{mat}} \quad ,$$

$$y_{pat} = b_{pat}\, x + e_{y_{pat}} \quad .$$

The estimates from these three equations are used to expand the LDSC-estimated S matrix containing genetic covariances, to also include variant-phenotype covariances (if the variant *x* is standardized, such that the *b* estimates are themselves standardized, then the *b* estimates are equal to the variant-phenotype covariances; if the variant *x* is not standardized, then multiplying the *b* estimate by the 2MAF(1-MAF), can be used to compute the corresponding variant-phenotype covariance). As described in Grotzinger et al.[5], the associated sampling covariance matrix of S is also expanded using cross-trait intercepts from LDSC in order to take any potential sample overlap (known or unknown) and/or shared stratification implied by the LDSC model into account. A model is then fit to the expanded S and V matrices that simultaneously estimates the terms in the Λ vector (using the minimal identification constraint $\lambda_{Direct} =$

1 in order to define the metric of F, as described earlier) and regresses F on the individual variant. This model is composed of the following two sets of equations.

$$Y = \Lambda \mathrm{F} + \mathrm{U} \quad ,$$

$$F = \gamma x + e \quad ,$$

where $\gamma$ is an unstandardized regression coefficient and $e$ is a residual. This model implies that

$$b_{direct} = \lambda_{direct} \cdot \gamma \quad ,$$

$$b_{mat} = \lambda_{mat} \cdot \gamma \quad ,$$

$$b_{pat} = \lambda_{pat} \cdot \gamma \quad .$$

Intuitively, the estimation of $\gamma$ can be conceived of as a WLS regression of the b effects from the three original GWAS on Λ, with the regression intercept fixed to 0, and reflects the meta-analytic estimate for the effect of the genetic variant on the target phenotype, disattenuating the direct GWAS, maternal GWAX, and paternal GWAX summary data based on the Λ terms, and weighting by the precision of their estimates. Thus, when the minimal identification constraint $\lambda_{direct} = 1$ is imposed, the WLS regression coefficient (which reflects the expected magnitude of b per 1 unit increase in $\lambda$ from 0) can be interpreted as the meta-analytic estimate of the three sets of summary statistics, scaled relative to the direct GWAS.

As explicated in section S3, the effective sample size for the direct GWAS and GWAX can be calculated as the observed sample size multiplied by the square of the corresponding attenuation factor ($\lambda$). Thus, under conditions of no sample overlap, the effective sample size for the summary statistics produced under the multivariate model introduced here, can be calculated as

$$N_{eff} = \lambda_{direct}^2 n_{direct} + \lambda_{mat}^2 n_{mat} + \lambda_{pat}^2 n_{pat} \quad .$$

For case-control traits, we may use this formula to calculate effective sample size for cases (substituting the corresponding number of cases for each constituent *n*), so as to obtain a sensible estimate of the effective proportion of cases in the sample when computing liability scale heritability.

### S7. The Heterogeneity Coefficient, $Q_{SNP}$

The model introduced in S5 allows for the possibility that each of the three sets of phenotypes (direct GWAS and two GWAX) is influenced by some genetic factors that are not shared across the three of them. This would be indicated in the empirical LSDC-estimated genetic correlation matrix (i.e. the standardized version of the S matrix) by off-diagonal elements less than 1.0, and in the model parameters as diagonal elements of Θ greater than 0. When this is the case, we expect to find some SNPs for which the regression coefficient on F does not well account for its pattern of associations with the three phenotypes. This will arise, for example, when a SNP has associations with only a subset of phenotypes, or the SNP has an association with some phenotypes incremental of its association with the general phenotype. For example, reports of parental AD history data may confuse other cognition-impairing disorders for AD at substantially higher rates than such disorders are mistaken for AD in the direct GWAS (which employ more rigorous diagnostic criteria to case ascertainment). In this example, genetic signal, e.g. for delirium, might be present in the GWAX summary data but not the direct GWAS data.

We can conceptualize the observed data as being generated by a model in which the total effect of a variant on a GWAS or GWAX indicator consists of a component that is mediated by the factor F, and (potentially) a component that occurs directly on the indicator and is unique of F, such that

$$b_{direct\ GWAS} = \lambda_{direct} \cdot \gamma \ \ (+\, b_{u\ direct})\ ,$$

$$b_{\ mat\ GWAX} = \lambda_{mat} \cdot \gamma \ \ (+\, b_{u\ mat})\ ,$$

$$b_{pat\ GWAX} = \lambda_{pat} \cdot \gamma \ \left(+\, b_{u\ pat}\right)\ ,$$

where the $\lambda s$ represent attenuation coefficient that reduces the expected effect size due to the indirect nature of the GWAX, and $b_u$s represent the unique effects of the variant on the individual indicators We cannot simultaneously estimate $\gamma$ along with all three b terms, as such a model is not locally identified (there are only 3 pieces of information regarding associations with the genetic variant, but we would like to estimate 4 such parameters). We therefore capitalize on the $Q_{SNP}$ statistic to identify variants displaying patterns of heterogeneity, i.e. variants whose empirical patterns of association with the three GWAS/GWAX phenotypes depart from expectations of the factor model. As detailed by Grotzinger et al.[5], and de la Fuente et al.[15], $Q_{SNP}$ is variant-specific chi squared distributed test statistic that indexes the difference in fit between the model described in section S6, in which the variant only affects the three indicators by way of its effect on F, and a model in which the variant directly effects the three indicators. We term the former model a common pathway model, and the latter model an independent pathways model. Significant $Q_{SNP}$ statistics indicate that expectations of the common pathway model are violated, and that the variant operates heterogeneously across the three indicators.
