## Supplementary Results for "Multivariate Modeling of Direct and Proxy GWAS Indicates Substantial Common Variant Heritability of Alzheimer’s Disease"

Javier de la Fuente,

Andrew D. Grotzinger, Riccardo E. Marioni, Michel G. Nivard,

& Elliot M. Tucker-Drob

### SR1. Meta-Analytic Results for Individual SNP Effects

We estimated meta-analytic summary statistics containing individual SNP effects on AD in our multivariate model using Genomic SEM, a Manhattan plot for which is provided in Figure S5, and qqplots for which are provided in Figure S6. The mean $\chi^2$ for the common factor GWAS output was 1.138. The mean $\chi^2(1)$ for $Q_{SNP}$ was 0.942, and there were no genome-wide significant ($p < 5 \times 10^{-8}$) hits for $Q_{SNP}$, indicating little evidence for genome-wide heterogeneity in SNP effects across direct GWAS, maternal GWAX, and paternal GWAX after the empirically derived attenuation coefficients ($\lambda$) are taken into account. The LDSC intercepts were all very close to 1.0, indicating that inflation of test statistics was predominately attributable to true polygenic signal rather than population stratification (Table 1).

We identified 280 independent significant SNPs and 93 lead SNPs in a total of 25 genome-wide significant loci associated with AD (Tables S4-S8). Of these, 23 significant loci were previously reported in published meta-analysis of GWAX and direct GWAS of AD by Marioni et al. and Jansen et al. and two were novel (genomic risk locus: 5, lead SNP: *rs114812713*, chromosome 6, $p = 1.12^{-11}$, nearest gene: OARD1; genomic risk locus: 10, lead SNP: *rs79832570*, chromosome: 8, $p = 4.50^{-8}$, nearest gene: SPATC1). Although not reported in either GWAS-GWAX meta-analysis genomic, locus 5 was reported in the direct GWAS by Kunkle et al. That we detected this locus using our multivariate method, in spite of it not having reach significant in the more traditional GWAS-GWAX meta-analyses, may be attributable to the previous meta-analyses not having used the optimal weights when combining GWAS and GWAX data. Moreover, risk locus 10 has not been previously reported in association with Alzheimer's disease, but it has been associated with hematological traits (e,g., eosinophil and neutrophil counts) and respiratory diseases.

For the genome-wide significant loci, we computed meta-analytic estimates using the inverse variance weighted approach and the Z approach. We applied each approach both naively (i.e. without correction) and with a correction for attenuation due to the indirect nature of the GWAX with the standard correction (Supplementary Table S6). As expected, based on the fact that the empirically derived $\lambda$ coefficients from our model were close to .5, we found that that the inverse variance weighted approach with the standard correction produced effect size estimates similar to those from our meta-analytic model. In contrast, the uncorrected approaches produced substantially deflated effect size estimates. The Z Statistic approach, even with the standard correction, still tended to produce somewhat deflated effect size estimates. This can be attributed to the fact that the correction employed corrected for attenuation due to the indirect nature for the GWAX, but did not correct for variability in prevalence rates stemming from the ascertained nature of the samples.

**SR2. Genetic Correlations with previous GWAS Meta-Analyses of Alzheimer's Disease and other External Correlates**

Figure S4 provides LDSC intercepts and LDSC-estimated genetic correlations of our multivariate AD meta-analysis with those of Marioni et al. and Jansen et al. with one another and with the direct GWAS of AD in IGAP and the GWAXs of maternal and paternal AD in UKB. It can be seen that the genetic correlations of the three meta analyses all exceed 1.0, indicating that the same genetic signal is tapped by each of them. The cross-trait intercepts are also very high (.67-.89) for the pairwise combinations of the three meta-analyses, as expected from their reliance on largely the same data. Investigating the genetic correlations between each of the three meta-analyses and the direct GWAS and two contributing GWAX, it can be seen that the Marioni and Jansen summary statistics demonstrate some highly out-of-bound associations (i.e. $r_g$ of 2.03 between Marioni meta analysis and IGAP; and $r_g$ of 1.81 and 1.74 between the Jansen meta-analyses and UKB maternal and paternal GWAX, respectively). In contrast, the associations between those produced by our multivariate method within Genomic SEM and the direct GWAS and two contributing GWAX are less extreme (e.g. the only out-of-bond estimated is the $r_g$ of 1.17 between the multivariate meta analysis and IGAP). It is possible that these differences stem from the differences in whether or not the optimal weights were used in the respective meta-analyses.

Figure S10 provides LDSC-estimated genetic correlations of our multivariate AD meta-analysis, and those by Marioni et al. and Jansen et al. with brain volume, educational attainment, and general cognitive function in the general population. AD risk, as indexed by Jansen et al. meta-analysis was more strongly genetically correlated with educational attainment ($r_g$=-.2) than was AD risk as indexed by the Marioni meta analysis ($r_g$=-.07) and the multivariate meta analysis ($r_g$=-.06). The three meta-analyses were more consistently related to a general genetic factor of cognitive function ($r_g$ = -0.19 for all three). There were no meaningful genetic associations with brain volume. Note that all LDSC analyses were based on common variants (MAF ≥ .01) outside of the MHC and APOE regions. Because other work has indicated very little evidence for genetic correlations between AD and other GWAS traits, we did not examine a wider range of genetic correlates.
