## Supplementary Figures for "Multivariate Modeling of Direct and Proxy GWAS Indicates Substantial Common Variant Heritability of Alzheimer’s Disease"

Javier de la Fuente,

Andrew D. Grotzinger, Riccardo E. Marioni, Michel G. Nivard,

& Elliot M. Tucker-Drob

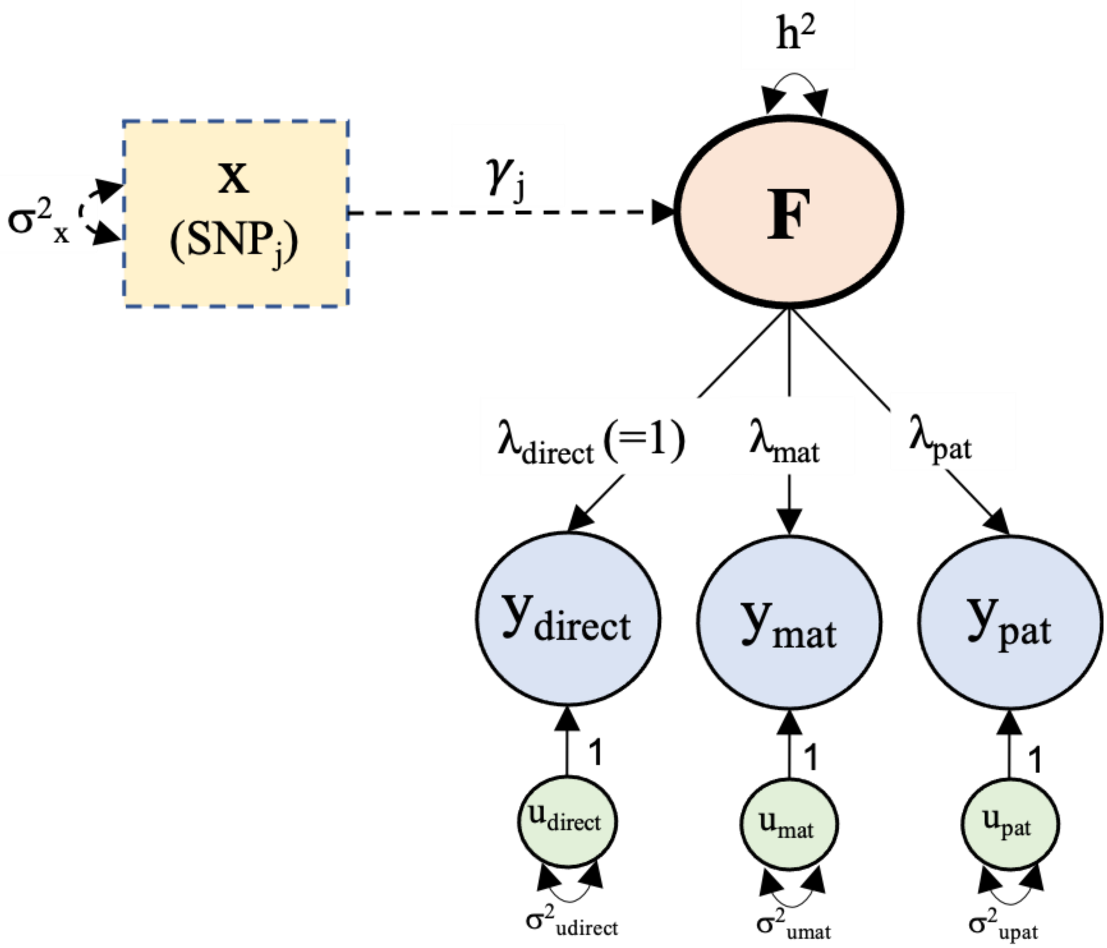


**Figure S1.** Path diagram for multivariate genetic analysis of Alzheimer's disease. We present the minimal identification constraint $\lambda_{direct} = 1$ such that the variance of the factor corresponds to the meta-analytic heritability estimate on scale of the direct GWAS. The dashed portion of the diagram represents the portion of the model that is specified to produce meta-analytic estimates for effects of individual SNPs.

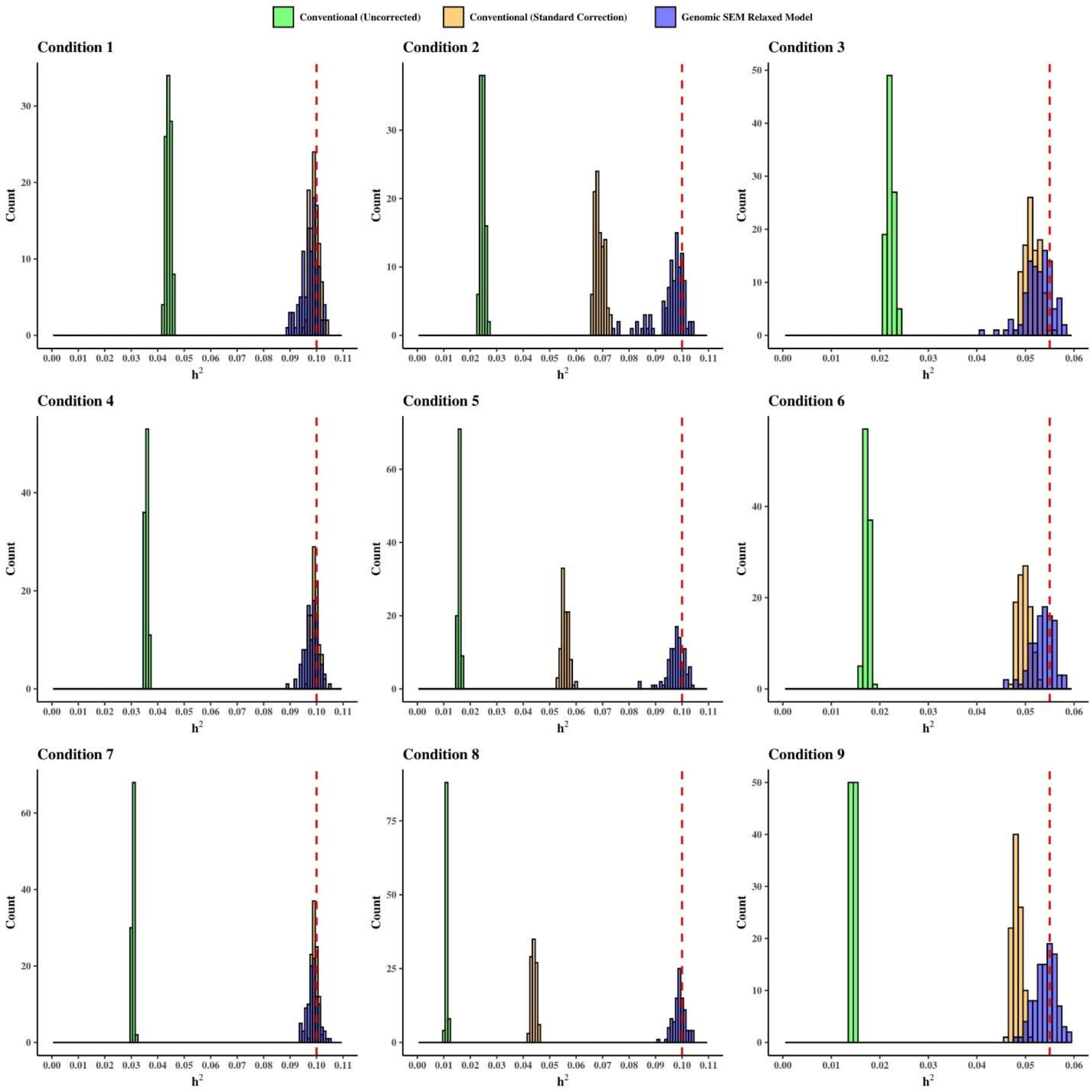


**Figure S2.** Simulation Results. Distribution of SNP heritability estimates ($h^2_{SNP}$) across simulation conditions.

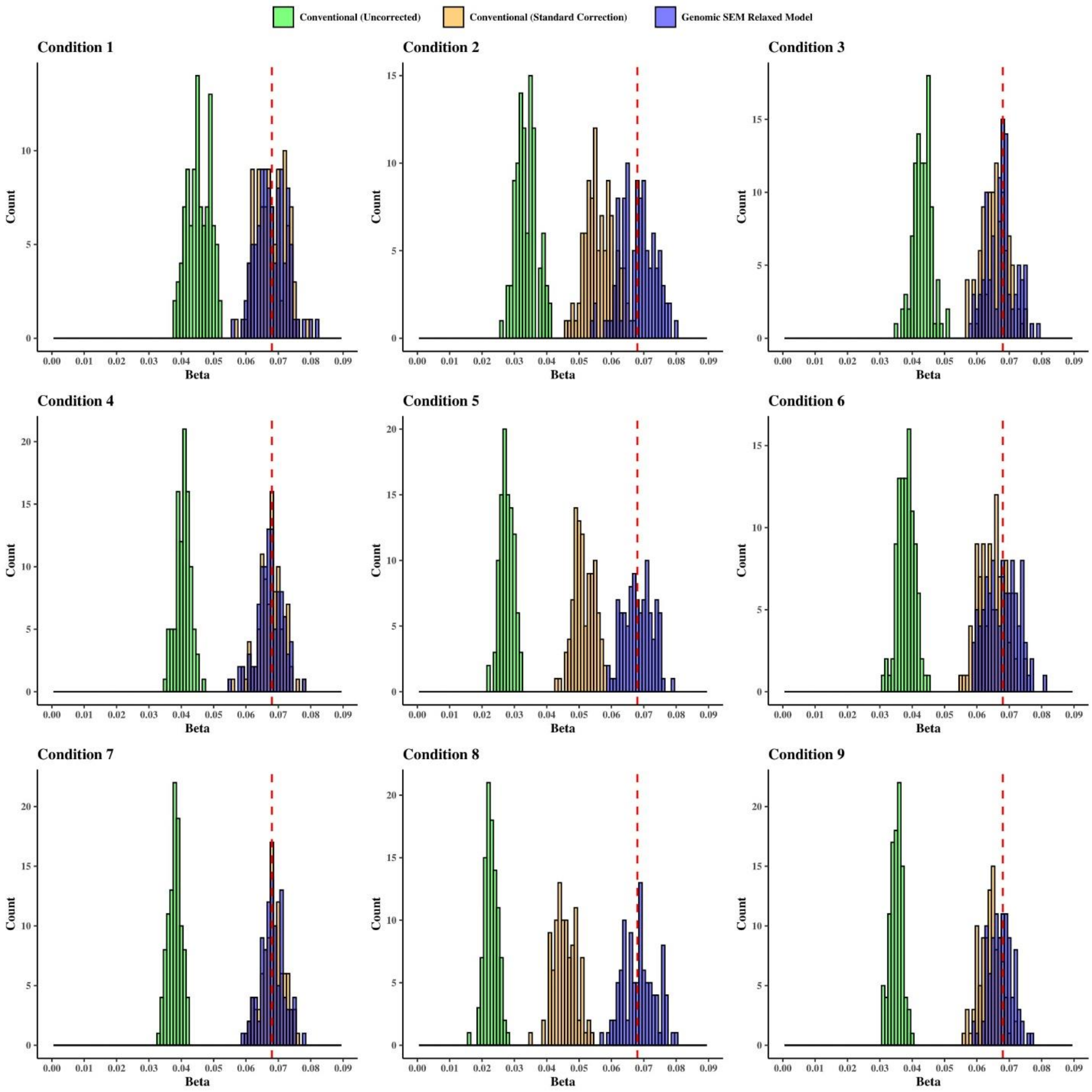


**Figure S3.** Simulation Results. Distribution of individual SNP effects across simulation conditions.

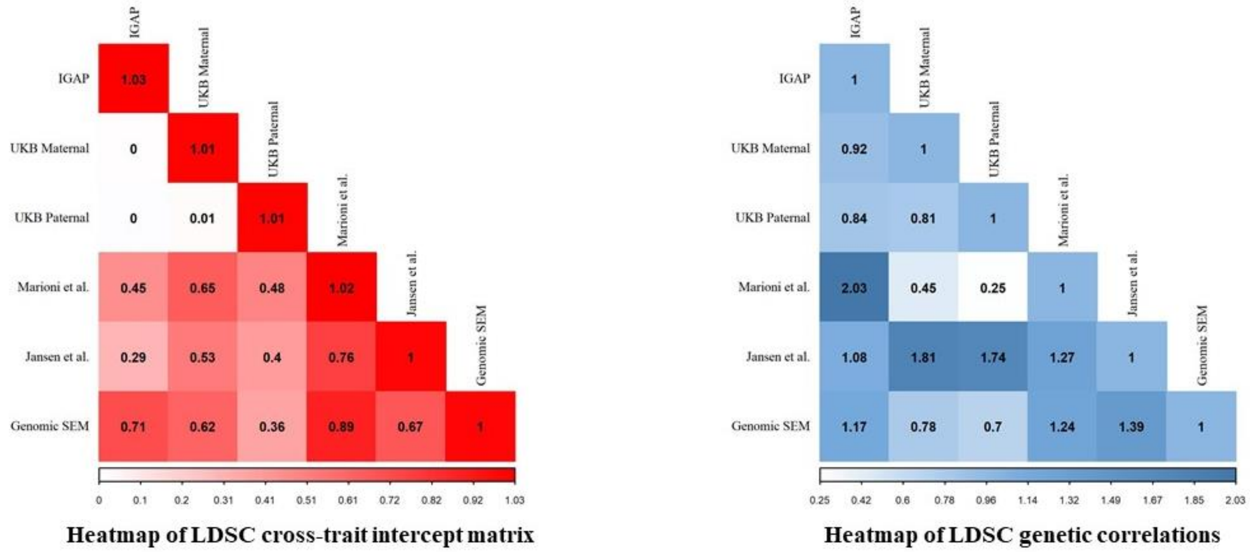


**Figure S4.** Heatmaps of LDSC cross-trait intercepts (left) and genetic correlations (right) among direct GWAS, maternal and paternal GWAX, and meta-analytic summary statistics of Alzheimer's disease from two previous studies, and from the Genomic SEM-based multivariate method introduced here.

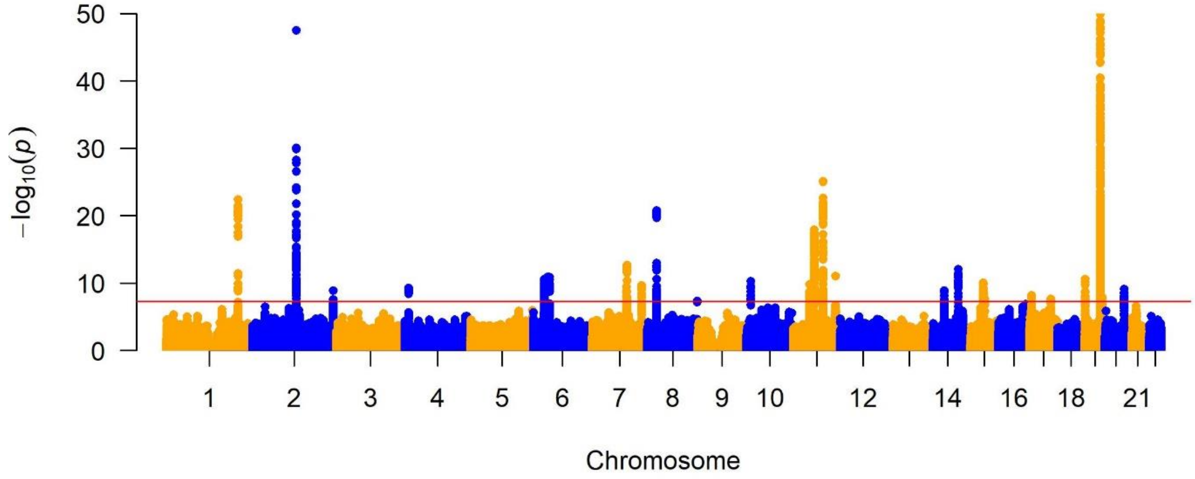


**Figure S5.** Manhattan plot for the common factor GWAS of Alzheimer's disease.

*Note:* the upper limit for the y axis was constrained to 50, the APOE region goes off-scale (lead SNP rs429358 -$\log_{10}$(p) = 308.653).

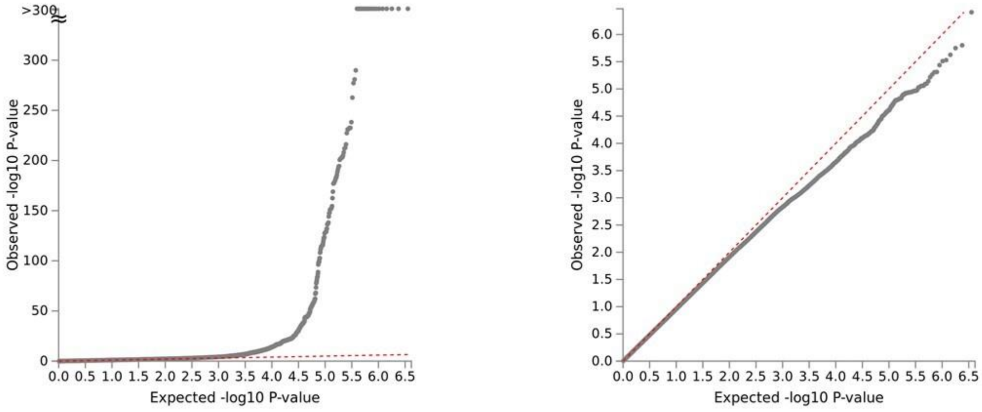


AD Genomic SEM Relaxed Model

Heterogeneity Q

Mean $\chi^2$ for AD GSEM = 1.138.
Mean $\chi^2$ for heterogeneity Q = 0.942

**Figure S6.** QQ-plot for AD Genomic SEM relaxed model and heterogeneity Q.

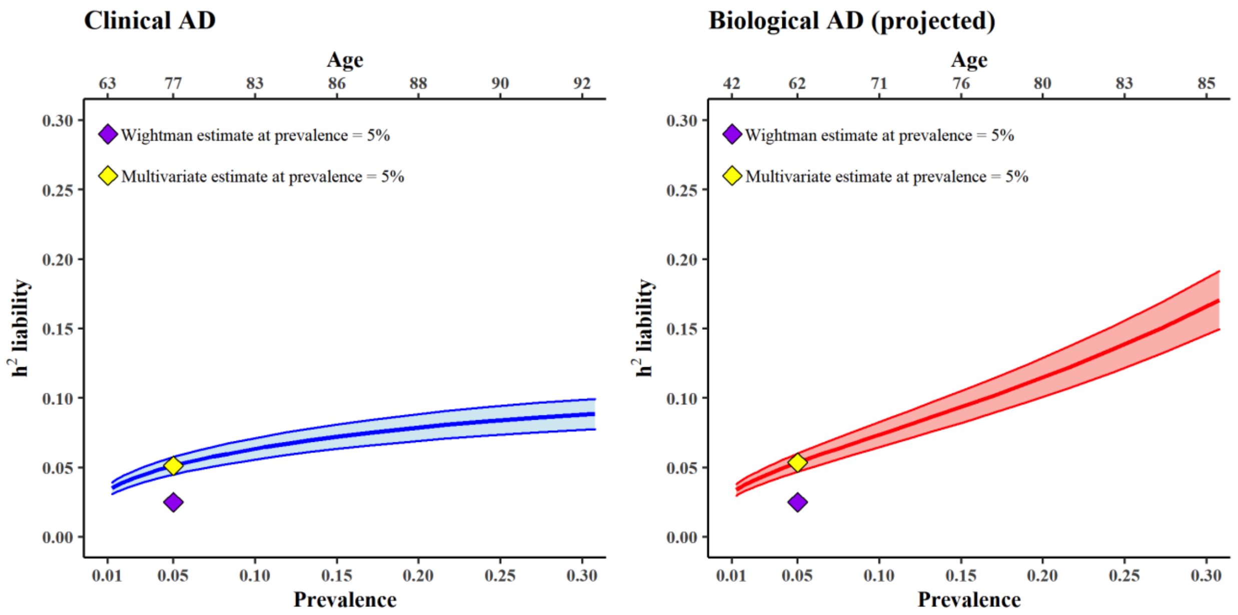


**Figure S7.** Estimated common variant SNP heritability of AD (outside of the MHC and *APOE* regions) according to different assumptions regarding the population prevalence of AD and biological AD, based on LDSC of summary statistics from our multivariate meta-analysis. We provide rough approximations of the age equivalences of each prevalence rate on the top x axis. The purple diamond represents the estimate of 2.5% by Wightman et al.(*4*), which was based on an assumed population prevalence rate of 5%. The yellow diamond represents the estimate from the multivariate model introduced here, using the same assumed population prevalence rate of 5% (Clinical AD $h^2$ = 0.051; Biological AD $h^2$ = 0.053). The shading area around the line reflects +/- 1 SE of the $h^2$ estimate. The steeper shift in the SNP heritability of AD for biological AD compared to clinical AD as a function of population prevalence stems from the correction for undetected biological AD within control participants who primarily only been screened for clinical AD. Thus, as the assumed prevalence rate of biological AD increases, the extent of case contamination in control participants increases, and the correction for undetected AD in control participants produces more dramatic increases in the projected heritability.

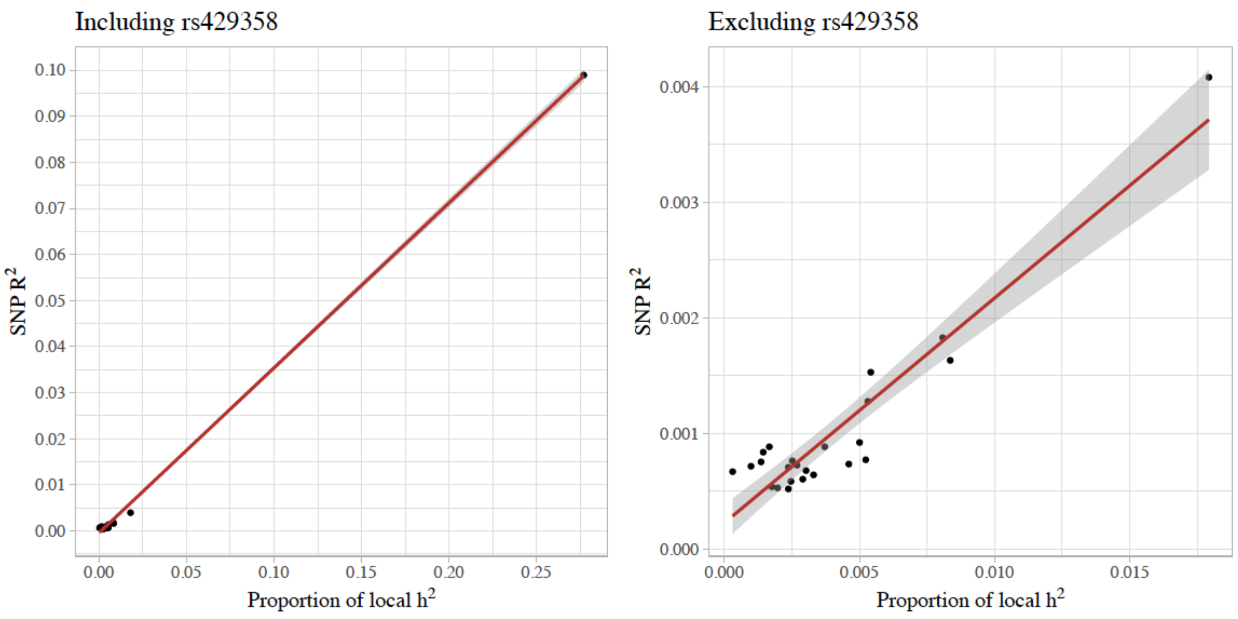


**Figure S8.** Association between local SNP heritability and GWAS effect sizes for GWAS loci that were genome wide significant in the multivariate GWAS including the APOE locus containing lead SNP rs429358 (left) and excluding this locus (right).

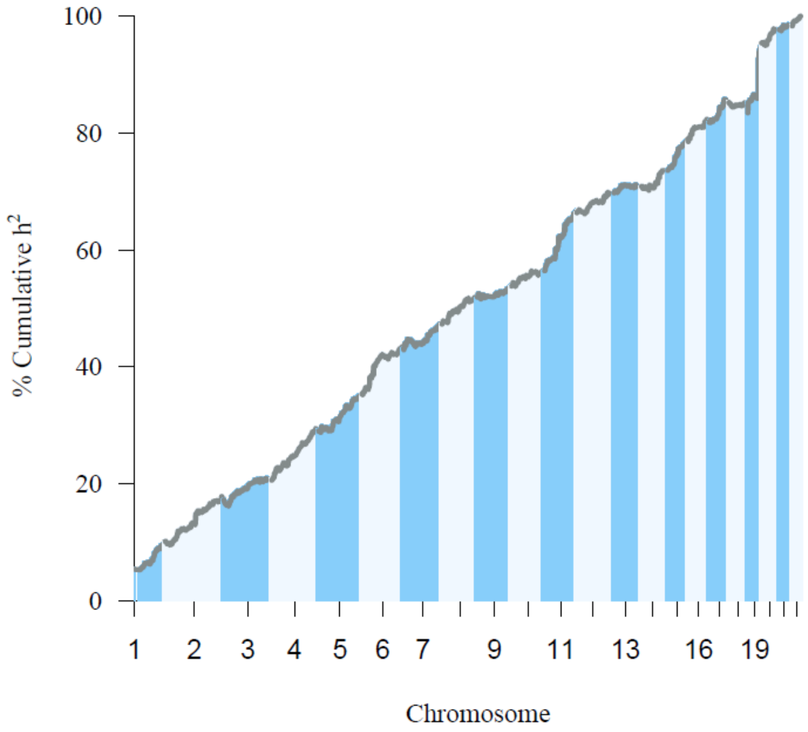


**Figure S9.** Local SNP heritability results for IGAP.

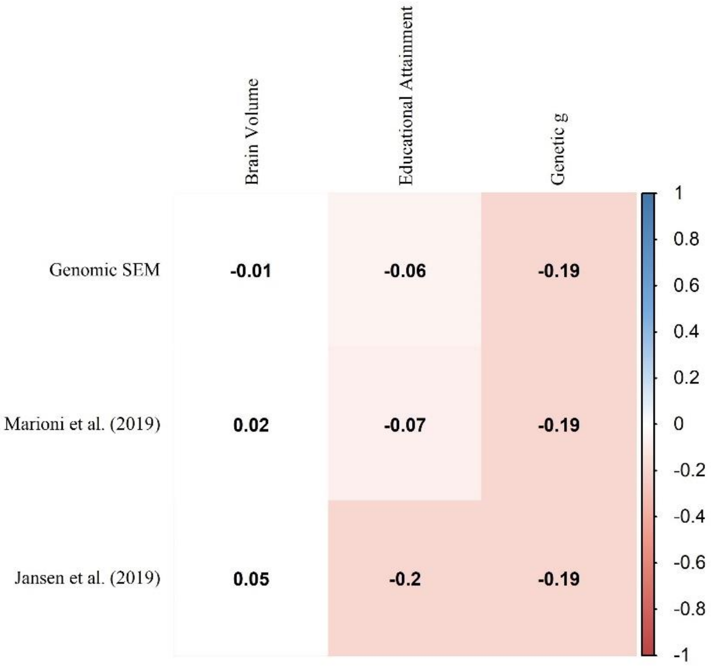


*Note: only significant correlations are colored ($p < 0.05$).*

**Figure S10.** Heatmap of LDSC genetic correlations among meta-analyses of case-control and proxy-phenotype family history of Alzheimer's disease and external correlates of Alzheimer's disease.
